# Reliability of biomechanical mensuration methods of the sagittal cervical spine on radiography used in clinical practice: A systematic review of literature

**DOI:** 10.1101/2025.05.06.25327135

**Authors:** Douglas F. Lightstone, Joseph W. Betz, Jason W. Haas, Paul A. Oakley, Joseph R. Farrentelli, Ibrahim M. Moustafa, Deed E. Harrison

## Abstract

Biomedical literature assessing reliability of mensuration of sagittal cervical spine alignment on radiographs is limited. This review aims to systematically identify and assess reliability studies on biomechanical assessments of the sagittal cervical spine used in clinical practice. The study design was registered with PROSPERO (CRD42023402990). Funding was from CBP, Non-Profit (Eagle, ID, USA). Inclusion criteria involved studies in English with: human subjects, radiography of the sagittal cervical spine, and reliability analysis on biomechanical mensuration of sagittal cervical spine. Exclusion criteria involved studies with: geometric modeling, animals, cadavers, phantom mannequins, and non-radiographic studies. Pubmed, CINAHL, AltHealth Watch, and Web of Science databases were searched from inception through January 24, 2023. The quality appraisal tool for studies of diagnostic reliability (QAREL) assessed bias risk. Results are presented following the synthesis without meta-analysis (SWiM) in systematic reviews guidelines. Scrutiny of inclusion criteria yielded 51 articles. Results are limited due to heterogeneity of the various mensuration methods and statistical analyses. The preponderance of evidence found good to excellent reliability. The results of this systematic review show that sagittal radiographic mensuration of cervical spine biomechanics and alignment is a reliable method for the diagnosis and management of spine conditions in clinical settings.

## Introduction

Cervical spine disorders, such as neck pain (NP), are major contributors to the Global Burden of Disease (GBD) and are the fourth largest contributor to disability worldwide^1^. GBD investigations have found that over 200 million people suffer from cervical spine disorders with a higher prevalence in females and up to 70% of the population suffering in their lifetime. Most NP, for example, has been reported as acute, but more that 50% of the population who suffer from NP have chronic episodes and frequently report dysfunction due to pain [1]. Patients suffering from chronic NP have reduced productivity and quality of life (QOL) and experience increased dysfunction and years lived with disability (YLD) with reports demonstrating 244 for every 100,000 [2].

Cervical spine disorders, including NP, have been studied at great length with regards to mechanisms of injury, genetic contributions, morphological abnormalities, psychosocial models, and biomechanics with limited conclusions and consensus on causes [3]. A recent emergence of bio-psychosocial models of cervical spine disorders have been popularized [4], but neglect structural and biomechanical contributions [5]. Understanding the comprehensive aspects of cervical spine disorders provide clinicians and patients with the best possible treatment recommendations. Further, studies have reported that neglecting biomechanical factors leads to poor quality outcomes [6].

Understanding the role of altered sagittal cervical spine alignment is of growing importance and interest in healthcare [7,8]. Sagittal spinal alignment has been studied since the beginnings of x-ray radiography [9]. Studies have found that normal, healthy sagittal cervical biomechanical measurements are desirable and important for improved clinical outcomes [10–14]. Extensive investigations show that alterations in the sagittal cervical lordosis have a significant impact on patient QOL and well-being, pain, and dysfunction and disability [15,16].

Valid, reliable methods of biomechanical mensuration of the cervical spine are essential for injury triage, diagnosis, and patient management (including treatment options or referral recommendations) patients with cervical spine disorders [15–18]. Cervical spine biomechanical measurements assist healthcare providers make the best clinical decisions in terms of treatment recommendations and continued management throughout the treatment process to assess patient progress and overall prognostics of cervical spine disorders [19–21]. The analysis of sagittal cervical configuration and lordosis are imperative in spinal surgery with the current techniques making use of vertebral segmental and global rotations and translations, sagittal balance, cranio-cervical measurements, and skull to thorax radiographic parameters [22]. Recent studies have shown that unhealthy sagittal cervical balance is a poor prognostic indicator for improvement post-surgery [23]. Sagittal cervical spinal mensuration and assessment is also necessary for conservative, spinal rehabilitation interventions such as those which require spinal therapeutic exercises and mechanical traction forces.

There are various methods of biomechanical analysis of cervical spinal alignment using radiography. That said, we found no systematic reviews of the literature (SROL) assessing the reliability of these methods using radiography. Given the significant impact of cervical spine disorders on GBD and YLD, the reliability of cervical spine radiographic biomechanical analysis would help to determine the best mensuration methods. The aim of this SROL is to:

1. Document the extent of the scientific literature reporting on the intra- and inter-examiner reliability of the biomechanical assessment of the sagittal cervical spine on radiography used in clinical practice.
2. Assess the quantitative and qualitative intra- and inter-examiner reliability of biomechanical mensuration methods for the sagittal cervical spine on radiography.
3. Appraise the bias risk and quality of the reliability studies.
4. Compare the reliability of the various biomechanical mensuration methods.
5. Document strengths and limitations of current scientific literature on the reliability studies and recommendations for future research on this topic.

Our hypothesis is that the available literature reporting on the on the intra- and inter-examiner reliability of the biomechanical assessment of the sagittal cervical spine on radiography used in clinical practice provides sufficient low bias risk with moderate to high-quality studies that establish good or better reliability.

## Methodology

Prior to the article search, the design of this SROL was registered with PROSPERO (CRD42023402990) and can be found by searching at https://www.crd.york.ac.uk/prospero/ in accordance with the standards of systematic review reporting, a PRISMA 2020 flow design, and statement [86]. PRISMA 2020 flow design and statement was initially created specifically for the effect of health interventions and encouraged multiple other SROL application to also follow this design. The use of PRISMA confirms the timeline of this review, the interpretation and presentation by the reviewers and the results that were found. This investigation, interpretation, presentation, and reportage were all in accordance with PRISMA checklists [24].

**Figure 1** demonstrates the preferred reporting items for systematic review and meta-analyses (PRISMA) flow diagram for selection, inclusion and exclusion used in this SROL.

**Figure 1.**
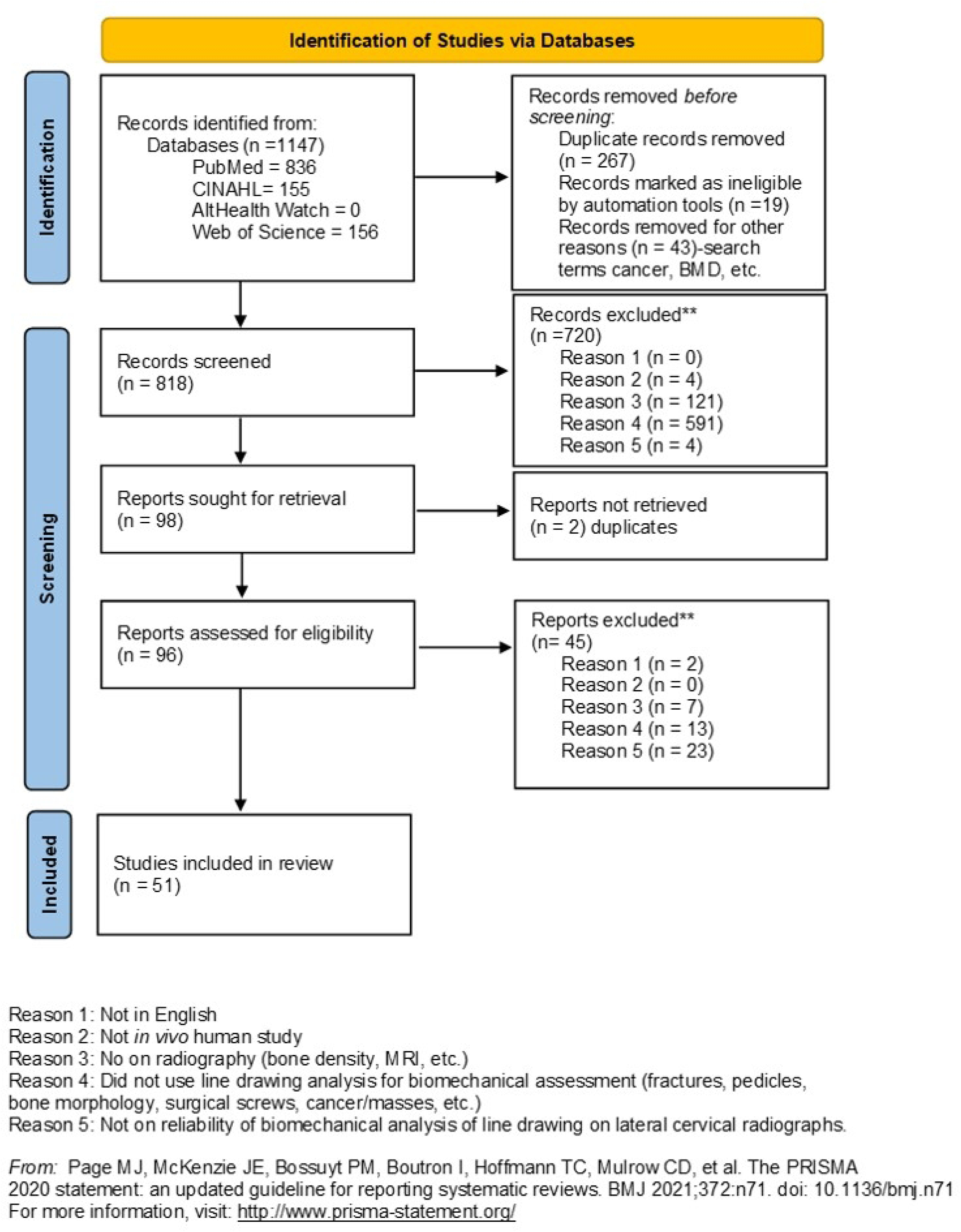
Preferred reporting items for systematic review and meta-analyses (PRISMA) flow diagram.

### Systematic Review Search Strategy

PRISMA checklists outline the search and interpretation process for this SROL. This strategy utilized a patient/population, intervention, comparison, and outcomes (PICO) alternate approach. The PICO alternate design was followed in accordance with search strategy for peer review of electronic search strategies (PRESS) checklist [25]. The librarian search databases consisted of PubMed, Alt Health Watch, CINAHL, and Web of Science (total databases) from the initial date of the database inception through January 24, 2023. The search MeSH terms studied by a master of library science are expanded in the Results and Figure 2.

**Figure 2.**
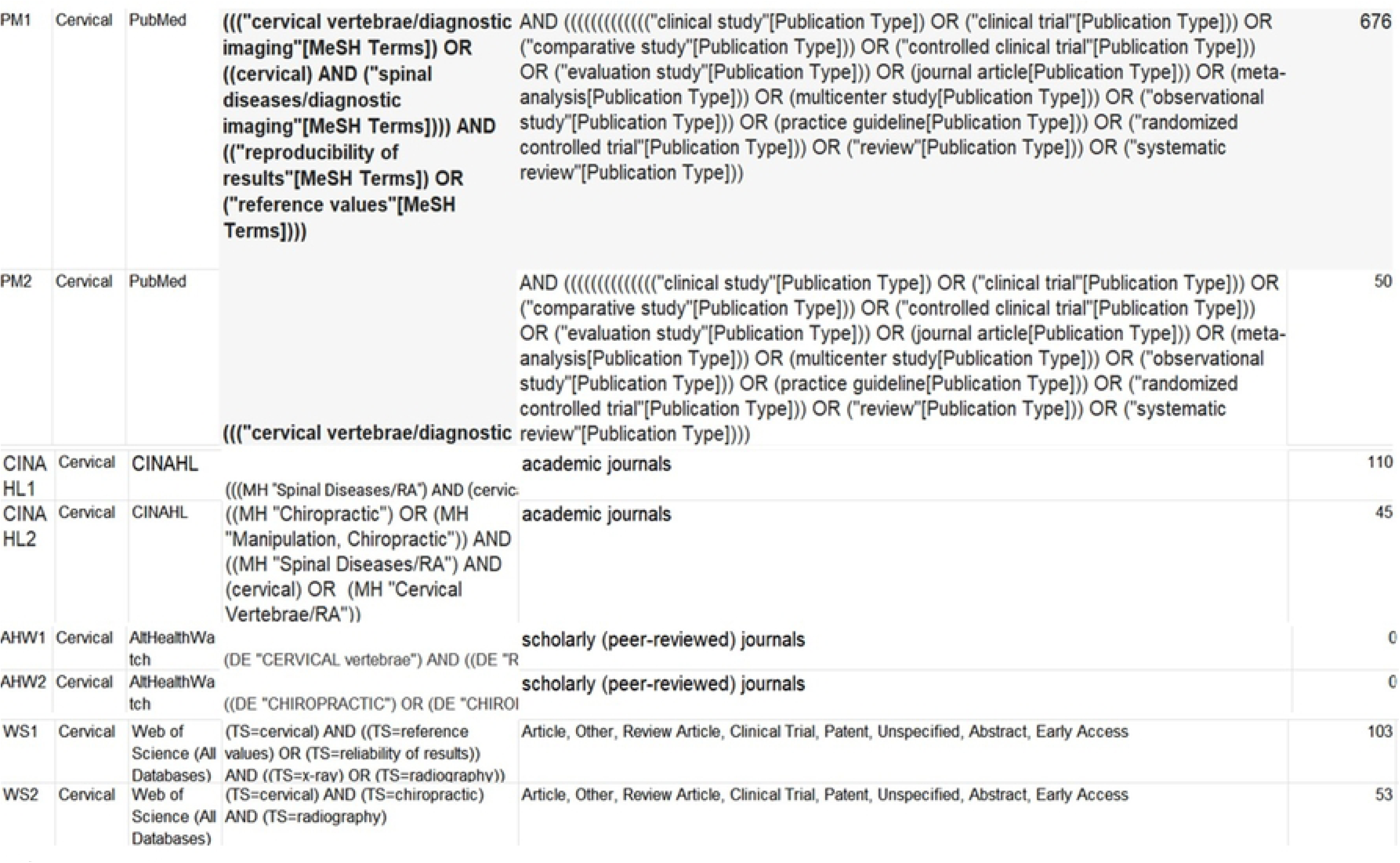
Expanded SROL search and MeSH terms performed by a master of library science.

### SROL Inclusion and Exclusion Criteria

The inclusion criteria for the search strategy for this SROL required the publications studied reliability of a biomechanical mensuration method of the sagittal cervical spine using radiography. Each study must have used radiography of the sagittal cervical spine on either asymptomatic or participants with defined cervical spine conditions and where the studies included biomechanical mensuration of sagittal cervical spine.

The exclusion criteria for this SROL were as follows:

- Criterion 1: The study was not in English. Note, a search was made for translations of manuscripts that were not in English and were not excluded if an English translation was found.
- Criterion 2: The study was not an *in vivo* human study.
- Criterion 3: The study was not about radiography.
- Criterion 4: The study was not about line drawing or mensuration for biomechanical analysis.
- Criterion 5: The study was not on the reliability of biomechanical analysis of line drawing on lateral cervical radiographs.

All non-radiographic studies and studies that did not perform radiographic biomechanical mensuration parameters of the lateral cervical spine were excluded from the final manuscript reviewal process. Since there are a multitude of methods for measuring cervical vertebral displacement, all measurement methods were included. Figure 3 depicts these methods.

**Figure 3.**
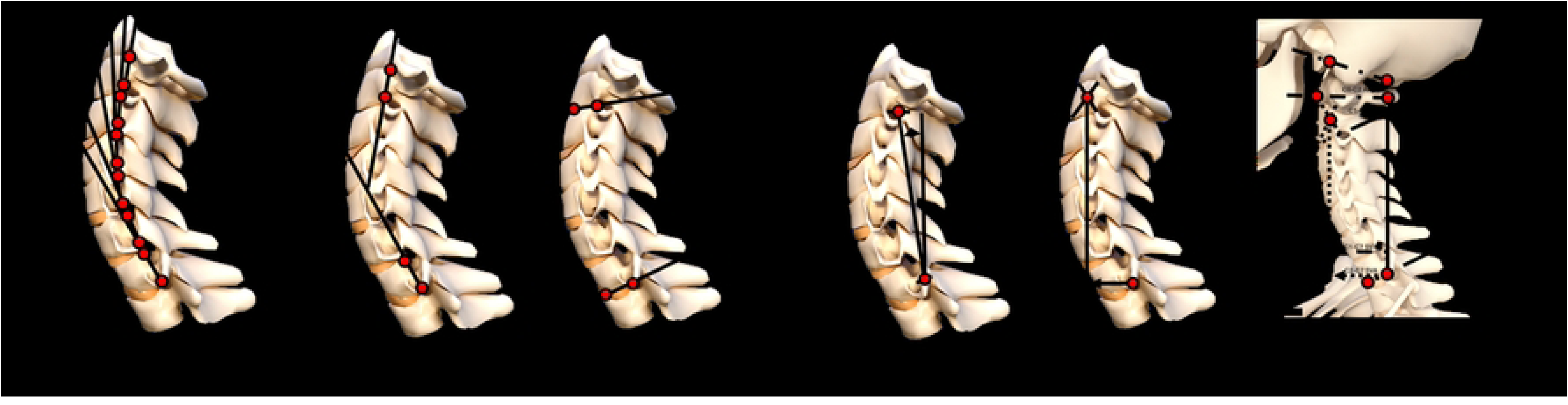
Examples of biomechanical mensuration methods of the sagittal cervical spine on radiography used in clinical practice. A) Harrison posterior tangent method, absolute rotation angle from C2 to C7 vertebrae (ARA C2-C7) and intersegmental relative rotational angles (RRA) from C2 to C7; B) Jackson physiological stress lines without angulation method for measuring C2-C7 curvature; C) Sagittal cervical Cobb angle C2-C7; D) Sagittal cervical K-line tilt; E) Sagittal vertical axis from C2 to C7 (SVA C2-C7); and F) CO-C2 angle, C1-C2 angle, sagittal cervical Cobb angle C2-C7, SVA C1-C7, SVA C2–C7.

### Selection of Studies for Systematic Review

The initial search strategy screened abstracts and titles for the inclusion and exclusion criteria outlined above. The use of automation tools allowed further screening of studies to determine if there were duplicates and there was found a difference between the original database and the results. Authors DFL and JWB screened all abstracts and studies based on the inclusion and exclusion criteria and discrepancies or disagreements were settled by a third reviewer (JWH) resulting in consensus. The articles that met inclusion criteria with no exclusion criteria were used in the formulation of the results of this SROL.

### Review Study Characteristics and Data Collection

In the final screening, authors DFL and JWB screened the full text articles, retrieved by the master of library science, based on the inclusion and exclusion criteria and discrepancies or disagreements were settled by a third reviewer (JWH) resulting in consensus. Authors JRF and DFL created spreadsheets and files for interpretation in Microsoft Excel (S1 Table). Information was extracted from the studies including author and year of study, sample number and demographic information, number of examiners, repeat analysis detail, types of imaging, method of analysis, intra- and/or inter-examiner reliability analysis(es), and SEM and/or MAD. No assumptions were made regarding missing or unclear information. Missing data was recorded as ‘N’ for no in data tables (Table 1). The studies were then assessed for risk of bias, study quality, intra- and inter-examiner reliability statistical analyses, and SEM and MAD (Figures 4 and 5, S2 and S3 Tables).

**Figure 4.**
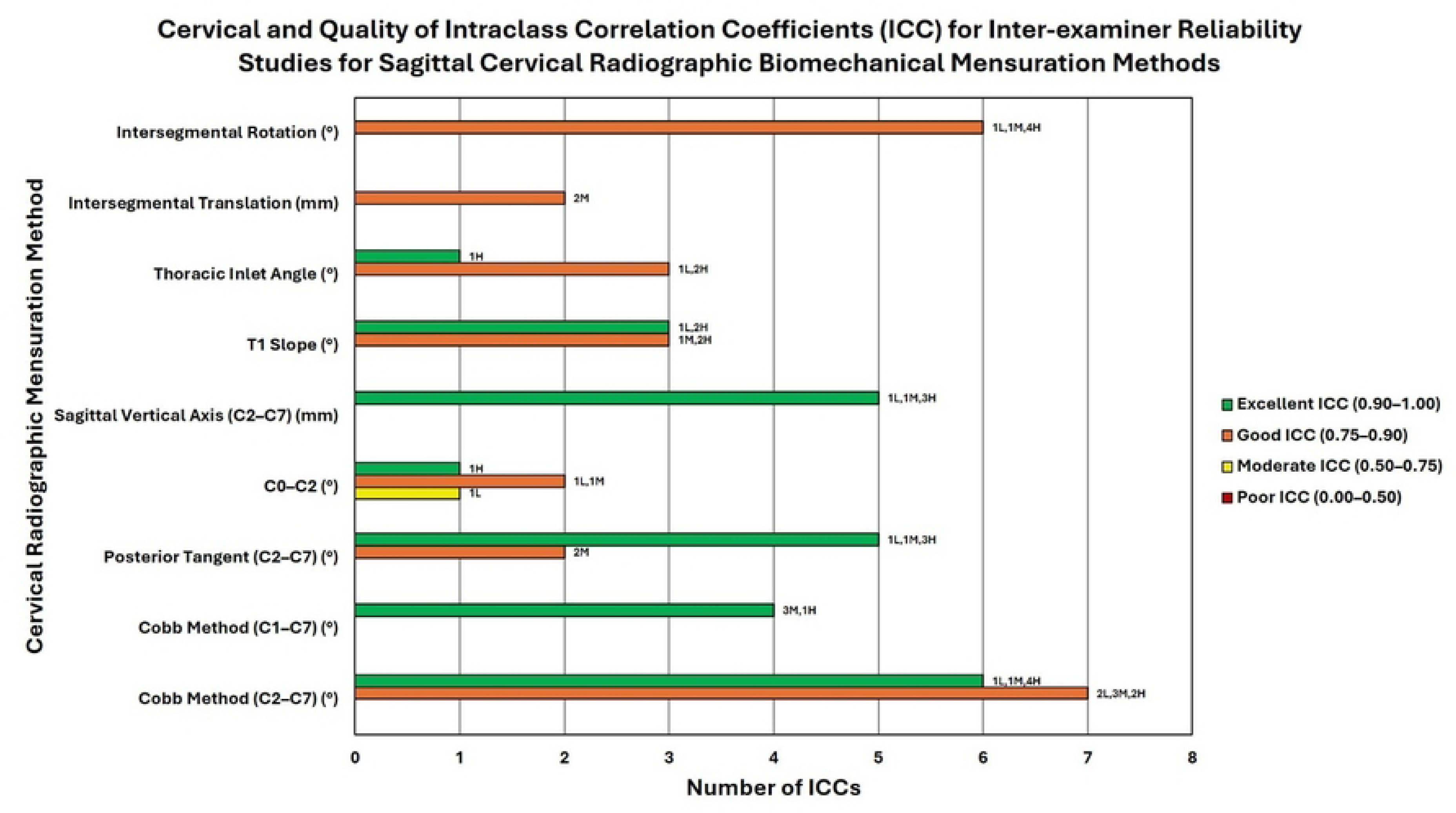
Cervical and quality of ICC of inter-examiner reliability studies for sagittal cervical radiographic biomechanical methods. This cluster bar chart represents the quality of reliability, quality of study, and number of studies found for inter-examiner reliability of sagittal cervical radiographic biomechanical mensuration methods. Each mensuration method (y-axis) may have up to four bars showing the number of ICCs (x-axis) for each cervical radiographic mensuration method (y-axis) representing the quality of ICC (excellent, good, moderate, or poor; see legend for color). The data labels at the outside end of the bars represent the number of low (L), moderate (M), and high (H) quality studies (as determined using the QAREL instrument) that comprise the total number of studies for a given ICC quality (excellent, good, moderate, or poor). For example, the Posterior Tangent Method (C2–C7) shows 8 excellent inter-examiner ICCs (comprised of 5 moderate-quality studies and 3 high-quality studies). The unit of measurement (° or mm) for each cervical radiographic biomechanical mensuration method is indicated in parentheses next to the mensuration method.

**Figure 5.**
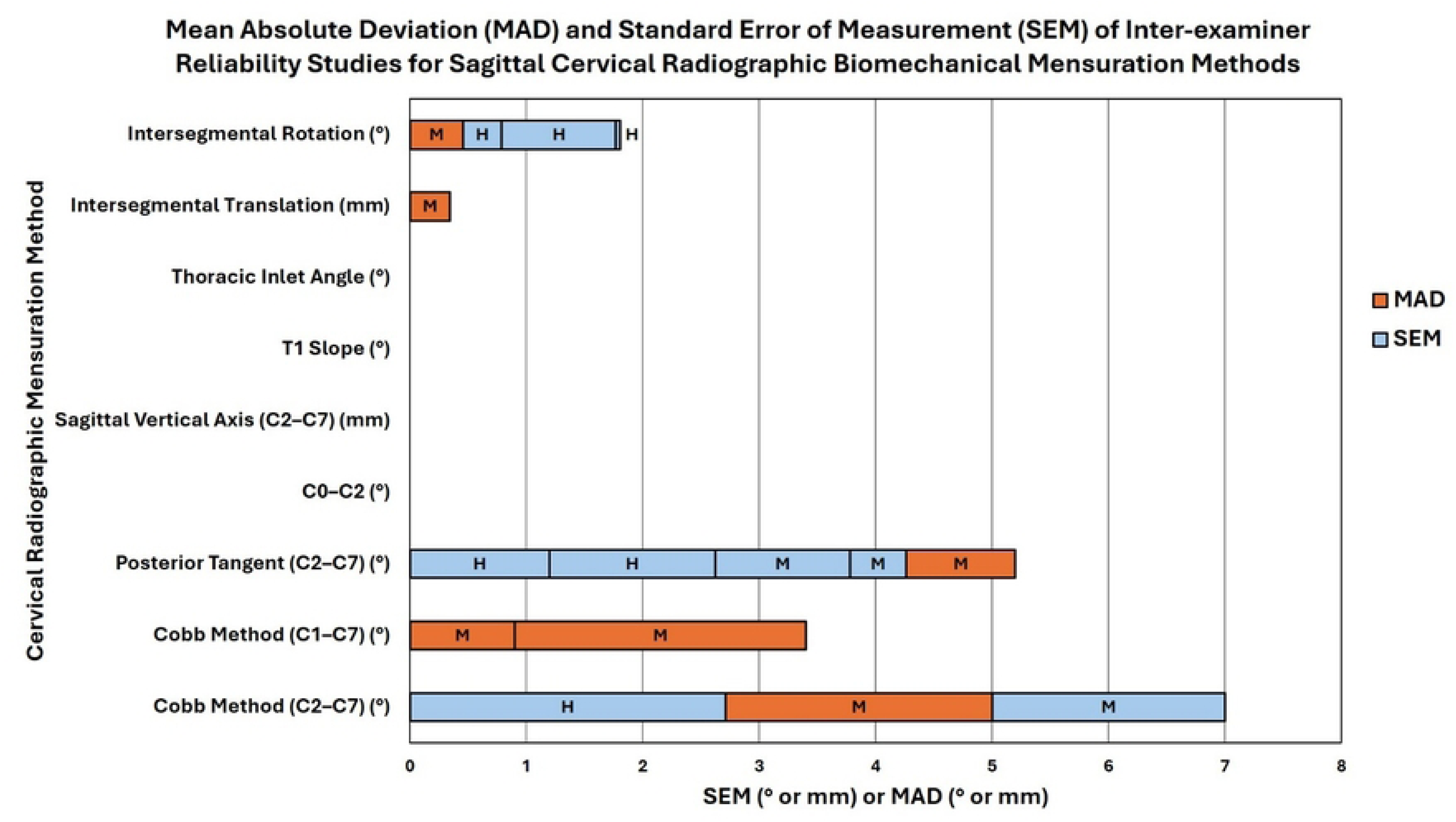
MAD and SEM of inter-examiner reliability studies for sagittal cervical radiographic biomechanical mensuration methods. This is a stacked bar chart representing the mean absolute deviation (MAD) and standard error of measurement (SEM) found in inter-examiner reliability studies for sagittal cervical radiographic biomechanical mensuration methods. The stacked bars show the number values of the MAD and SEM (x-axis; see legend for color) for each cervical radiographic mensuration method (y-axis) and the letter in each bar represents the quality of the study as determined by the QAREL instrument (L = low-quality, M = moderate-quality, and H = high-quality). The unit of measurement (° or mm) for each cervical radiographic biomechanical mensuration method is indicated in parentheses next to the mensuration method.

**Figure 6.**
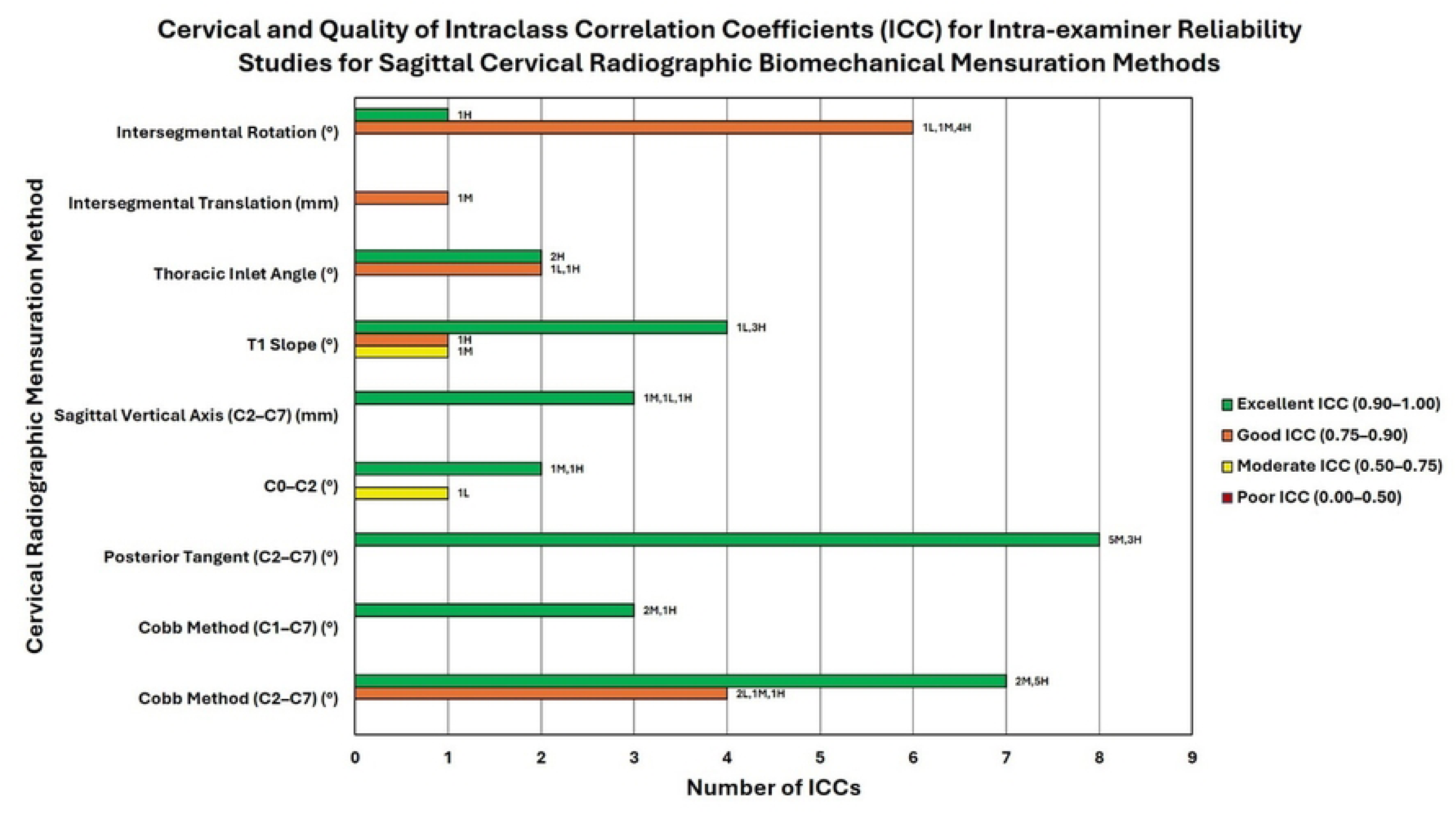
Cervical and quality of ICC of intra-examiner reliability studies for sagittal cervical radiographic biomechanical methods. This cluster bar chart represents the quality of reliability, quality of study, and number of studies found for intra-examiner reliability of sagittal cervical radiographic biomechanical mensuration methods. Each mensuration method (y-axis) may have up to four bars showing the number of ICCs (x-axis) for each cervical radiographic mensuration method (y-axis) representing the quality of ICC (excellent, good, moderate, or poor; see legend for color). The data labels at the outside end of the bars represent the number of low (L), moderate (M), and high (H) quality studies (as determined using the QAREL instrument) that comprise the total number of studies for a given ICC quality (excellent, good, moderate, or poor). For example, the Posterior Tangent Method (C2–C7) shows 8 excellent intra-examiner ICCs (comprised of 5 moderate-quality studies and 3 high-quality studies). The unit of measurement (° or mm) for each cervical radiographic biomechanical mensuration method is indicated in parentheses next to the mensuration method.

**Table 1.**
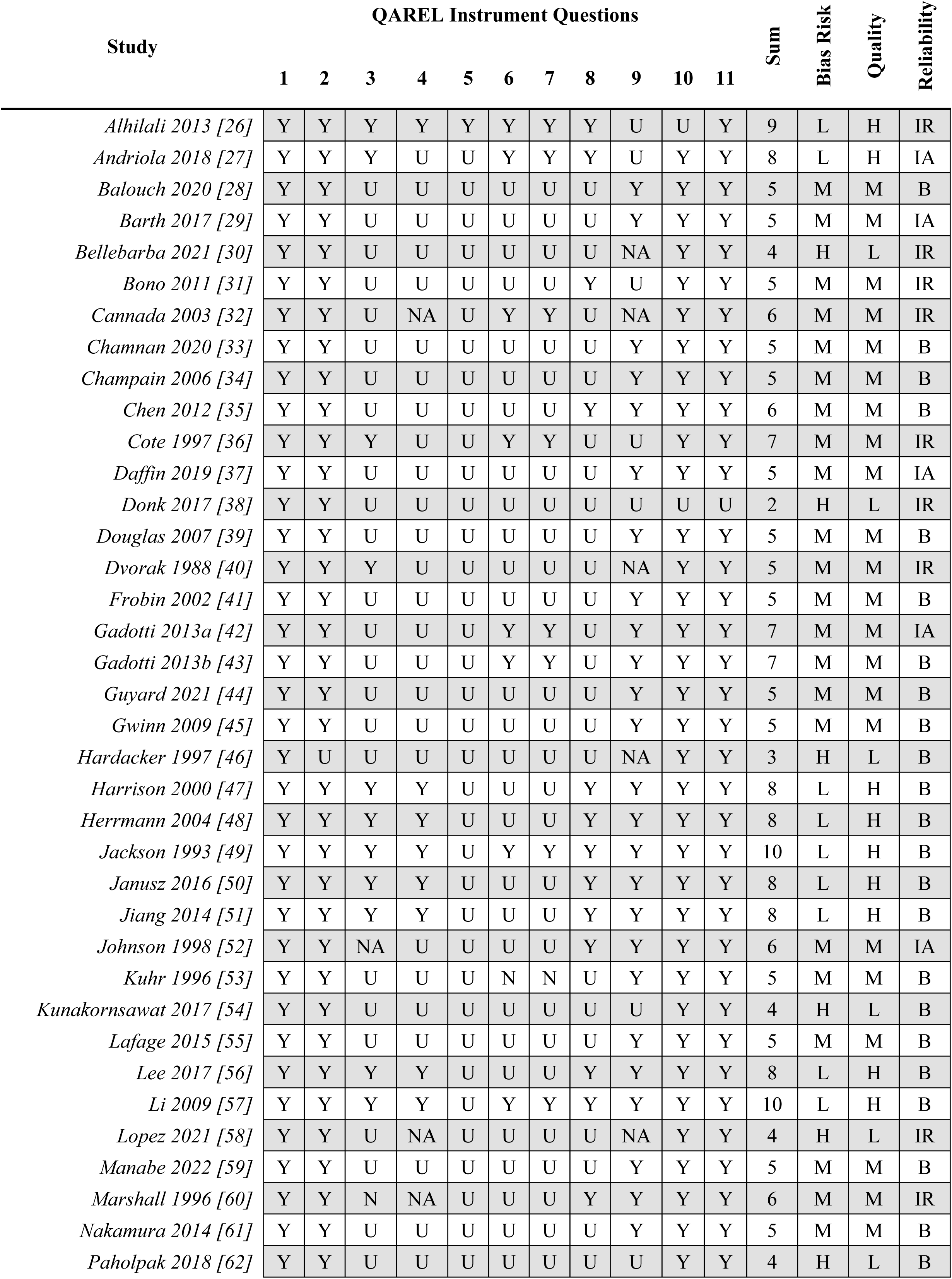
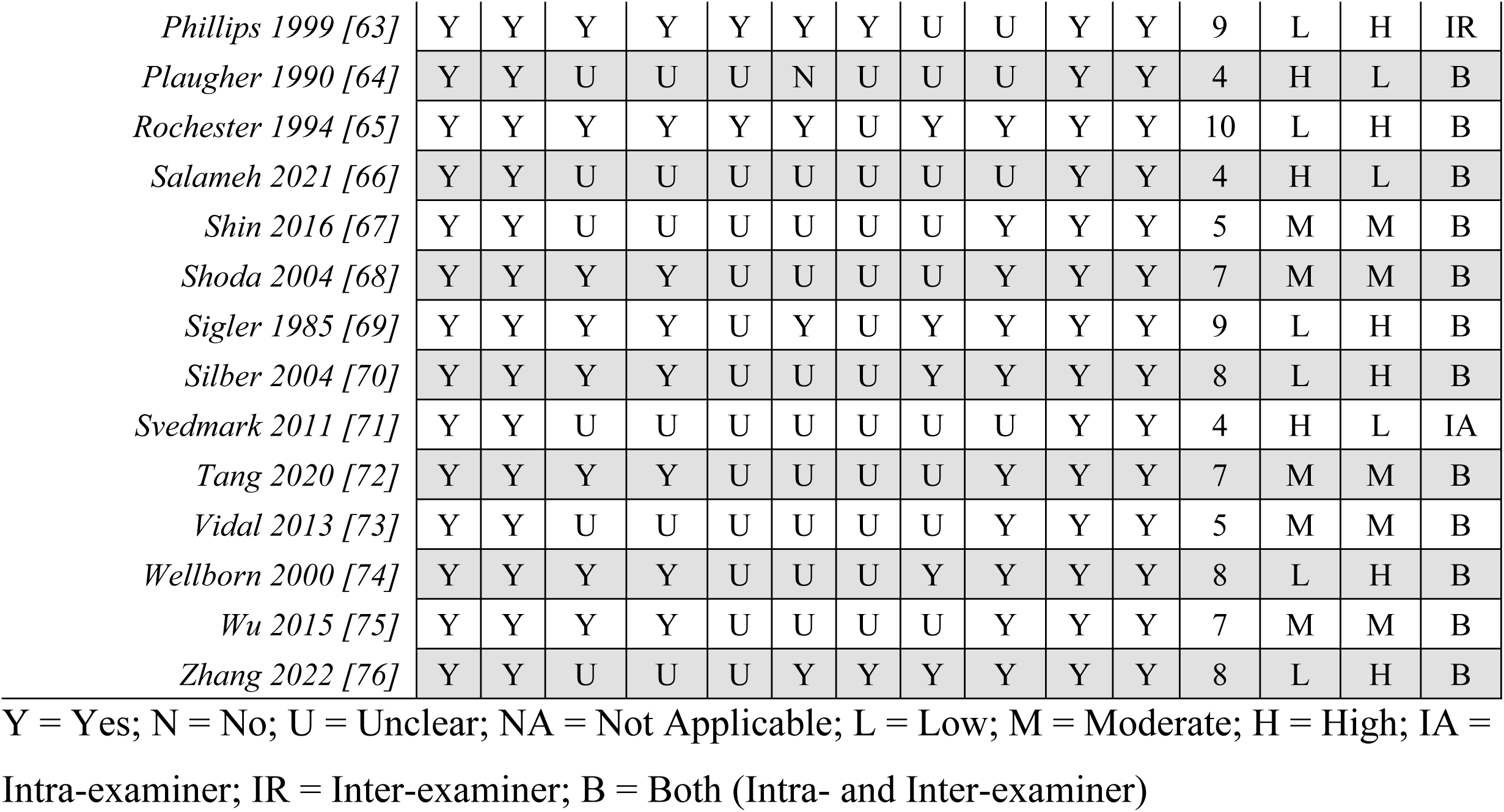
QAREL instrument results for sagittal cervical radiographic biomechanical mensuration intra- and inter-examiner reliability studies [26–76].

| Study | QAREL Instrument Questions |  |  |  |  |  |  |  |  |  |  | Sum | Bias Risk | Quality | Reliability |
| --- | --- | --- | --- | --- | --- | --- | --- | --- | --- | --- | --- | --- | --- | --- | --- |
|  | 1 | 2 | 3 | 4 | 5 | 6 | 7 | 8 | 9 | 10 | 11 |  |  |  |  |
| *Alhilali 2013 [26]* | Y | Y | Y | Y | Y | Y | Y | Y | U | U | Y | 9 | L | H | IR |
| *Andriola 2018 [27]* | Y | Y | Y | U | U | Y | Y | Y | U | Y | Y | 8 | L | H | IA |
| *Balouch 2020 [28]* | Y | Y | U | U | U | U | U | U | Y | Y | Y | 5 | M | M | B |
| *Barth 2017 [29]* | Y | Y | U | U | U | U | U | U | Y | Y | Y | 5 | M | M | IA |
| *Bellebarba 2021 [30]* | Y | Y | U | U | U | U | U | U | NA | Y | Y | 4 | H | L | IR |
| *Bono 2011 [31]* | Y | Y | U | U | U | U | U | Y | U | Y | Y | 5 | M | M | IR |
| *Cannada 2003 [32]* | Y | Y | U | NA | U | Y | Y | U | NA | Y | Y | 6 | M | M | IR |
| *Chamnan 2020 [33]* | Y | Y | U | U | U | U | U | U | Y | Y | Y | 5 | M | M | B |
| *Champaign 2006 [34]* | Y | Y | U | U | U | U | U | U | Y | Y | Y | 5 | M | M | B |
| *Chen 2012 [35]* | Y | Y | U | U | U | U | U | Y | Y | Y | Y | 6 | M | M | B |
| *Cote 1997 [36]* | Y | Y | Y | U | U | Y | Y | U | U | Y | Y | 7 | M | M | IR |
| *Daffin 2019 [37]* | Y | Y | U | U | U | U | U | U | Y | Y | Y | 5 | M | M | IA |
| *Donk 2017 [38]* | Y | Y | U | U | U | U | U | U | U | U | U | 2 | H | L | IR |
| *Douglas 2007 [39]* | Y | Y | U | U | U | U | U | U | Y | Y | Y | 5 | M | M | B |
| *Dvorak 1988 [40]* | Y | Y | Y | U | U | U | U | U | NA | Y | Y | 5 | M | M | IR |
| *Frobin 2002 [41]* | Y | Y | U | U | U | U | U | U | Y | Y | Y | 5 | M | M | B |
| *Gadotti 2013a [42]* | Y | Y | U | U | U | Y | Y | U | Y | Y | Y | 7 | M | M | IA |
| *Gadotti 2013b [43]* | Y | Y | U | U | U | Y | Y | U | Y | Y | Y | 7 | M | M | B |
| *Guyard 2021 [44]* | Y | Y | U | U | U | U | U | U | Y | Y | Y | 5 | M | M | B |
| *Gwinn 2009 [45]* | Y | Y | U | U | U | U | U | U | Y | Y | Y | 5 | M | M | B |
| *Hardacker 1997 [46]* | Y | U | U | U | U | U | U | U | NA | Y | Y | 3 | H | L | B |
| *Harrison 2000 [47]* | Y | Y | Y | Y | U | U | U | Y | Y | Y | Y | 8 | L | H | B |
| *Herrmann 2004 [48]* | Y | Y | Y | Y | U | U | U | Y | Y | Y | Y | 8 | L | H | B |
| *Jackson 1993 [49]* | Y | Y | Y | Y | U | Y | Y | Y | Y | Y | Y | 10 | L | H | B |
| *Janusz 2016 [50]* | Y | Y | Y | Y | U | U | U | Y | Y | Y | Y | 8 | L | H | B |
| *Jiang 2014 [51]* | Y | Y | Y | Y | U | U | U | Y | Y | Y | Y | 8 | L | H | B |
| *Johnson 1998 [52]* | Y | Y | NA | U | U | U | U | Y | Y | Y | Y | 6 | M | M | IA |
| *Kuhr 1996 [53]* | Y | Y | U | U | U | N | N | U | Y | Y | Y | 5 | M | M | B |
| *Kunakornsawat 2017 [54]* | Y | Y | U | U | U | U | U | U | U | Y | Y | 4 | H | L | B |
| *Lafage 2015 [55]* | Y | Y | U | U | U | U | U | U | Y | Y | Y | 5 | M | M | B |
| *Lee 2017 [56]* | Y | Y | Y | Y | U | U | U | Y | Y | Y | Y | 8 | L | H | B |
| *Li 2009 [57]* | Y | Y | Y | Y | U | Y | Y | Y | Y | Y | Y | 10 | L | H | B |
| *Lopez 2021 [58]* | Y | Y | U | NA | U | U | U | U | NA | Y | Y | 4 | H | L | IR |
| *Manabe 2022 [59]* | Y | Y | U | U | U | U | U | U | Y | Y | Y | 5 | M | M | B |
| *Marshall 1996 [60]* | Y | Y | N | NA | U | U | U | Y | Y | Y | Y | 6 | M | M | IR |
| *Nakamura 2014 [61]* | Y | Y | U | U | U | U | U | U | Y | Y | Y | 5 | M | M | B |
| *Paholpak 2018 [62]* | Y | Y | U | U | U | U | U | U | U | Y | Y | 4 | H | L | B |
| *Phillips 1999 [63]* | Y | Y | Y | Y | Y | Y | Y | U | U | Y | Y | 9 | L | H | IR |
| *Plaughner 1990 [64]* | Y | Y | U | U | U | N | U | U | U | Y | Y | 4 | H | L | B |
| *Rochester 1994 [65]* | Y | Y | Y | Y | Y | Y | U | Y | Y | Y | Y | 10 | L | H | B |
| *Salameh 2021 [66]* | Y | Y | U | U | U | U | U | U | U | Y | Y | 4 | H | L | B |
| *Shin 2016 [67]* | Y | Y | U | U | U | U | U | U | Y | Y | Y | 5 | M | M | B |
| *Shoda 2004 [68]* | Y | Y | Y | Y | U | U | U | U | Y | Y | Y | 7 | M | M | B |
| *Sigler 1985 [69]* | Y | Y | Y | Y | U | Y | U | Y | Y | Y | Y | 9 | L | H | B |
| *Silber 2004 [70]* | Y | Y | Y | Y | U | U | U | Y | Y | Y | Y | 8 | L | H | B |
| *Svedmark 2011 [71]* | Y | Y | U | U | U | U | U | U | U | Y | Y | 4 | H | L | IA |
| *Tang 2020 [72]* | Y | Y | Y | Y | U | U | U | U | Y | Y | Y | 7 | M | M | B |
| *Vidal 2013 [73]* | Y | Y | U | U | U | U | U | U | Y | Y | Y | 5 | M | M | B |
| *Wellborn 2000 [74]* | Y | Y | Y | Y | U | U | U | Y | Y | Y | Y | 8 | L | H | B |
| *Wu 2015 [75]* | Y | Y | Y | Y | U | U | U | U | Y | Y | Y | 7 | M | M | B |
| *Zhang 2022 [76]* | Y | Y | U | U | U | Y | Y | Y | Y | Y | Y | 8 | L | H | B |

Y = Yes; N = No; U = Unclear; NA = Not Applicable; L = Low; M = Moderate; H = High; IA = Intra-examiner; IR = Inter-examiner; B = Both (Intra- and Inter-examiner)

All statistical analyses for intra- and / or inter-examiner reliability, and the results of any statistical analysis were recorded (Figures 4-7, S2 and S3 Tables). Statistical analytical methods for reliability assessments included intra- and inter-class ICCs, Pearson correlation coefficients, Cohen’s kappa agreement, and Bland-Altman plots; all of which can have different reliability assessments and interpretations [77].

**Figure 7.**
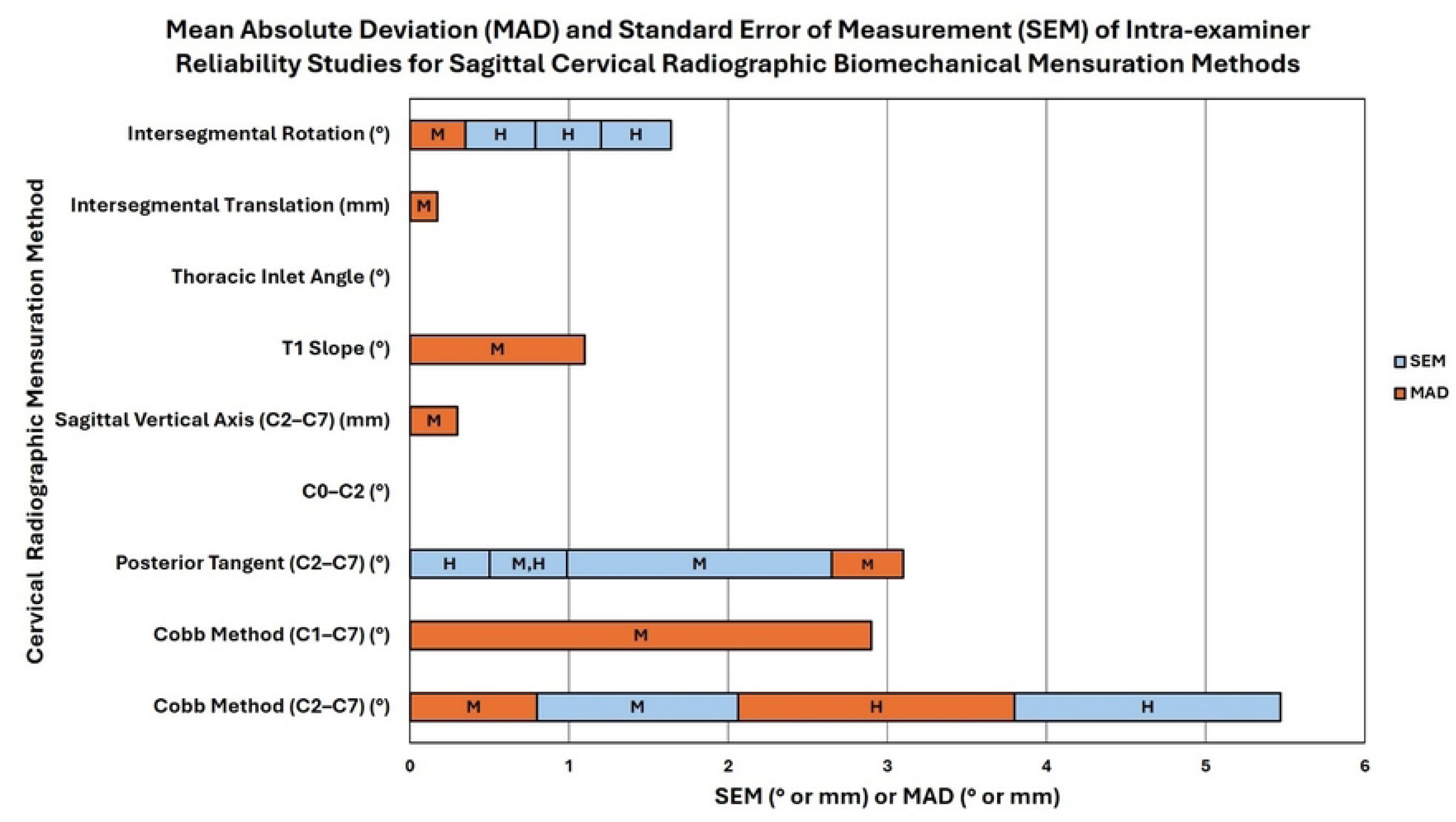
MAD and SEM of intra-examiner reliability studies for sagittal cervical radiographic biomechanical mensuration methods. This is a stacked bar chart representing the mean absolute deviation (MAD) and standard error of measurement (SEM) found in intra-examiner reliability studies for sagittal cervical radiographic biomechanical mensuration methods. The stacked bars show the number values of the MAD and SEM (x-axis; see legend for color) for each cervical radiographic mensuration method (y-axis) and the letter in each bar represents the quality of the study as determined by the QAREL instrument (L = low-quality, M = moderate-quality, and H = high-quality). The unit of measurement (° or mm) for each cervical radiographic biomechanical mensuration method is indicated in parentheses next to the mensuration method.

### Evaluation for Study Quality Assessment and Review Study Bias Assessment

The QAREL instrument used in reliability studies was confirmed and approved for this study by the authors. The assessment, described by Alfuth [78] and Konieczka et al. [79], was used to provide assessment of risk bias and study quality [80]. Following the assessment of the two initial reviewers (JRF and DFL) for any bias assessment of the studies, the third reviewer (JWH), confirmed bias risk and re-assessed confirmation of inclusion criteria. Utilization of this multi-step process for review of the articles originally acquired allows for significantly more critical appraisal of the data collected and offers much greater clinical application of the findings than selective, snap, or limited reviews.

### Conglomerated Study Synthesis and Data Analysis

The three-reviewer process incorporating the two initial assessments of data followed by the third referee review was used to reduce bias in data collection and improve systematic review quality and credibility. The three-reviewer process solved discrepancies, including missing data or missed duplication errors. It has been shown from prior studies that the use of multiple reviewers increases the quality and accuracy of data collection and synthesis in a SROL [81].

Prior studies have recommended the use of SWiM guidelines for SROL subgroup analysis to improve confidence and certainty of conclusions and findings [82].

## Results

### Systematic Review Selection Process and Search Terms

The discovery process yielded a total of 1147 potential articles. A deduplication tool in EndNote [83] was used for duplicate articles. Ineligible articles using automation were removed. Anomalous, pathological, and other conditions resulted in article removal. This resulted in 818 articles after filtering titles and abstracts for the following medical subject heading (MeSH) terms:

Search terms for PubMed were: “Cervical vertebrae/diagnostic imaging” (Medical Subject Headings (MeSH) Terms) OR “Cervical” AND “Spinal diseases/diagnostic imagining” (MeSH Terms) AND “reproducibility of results” (MeSH Terms) OR “reference values” (MeSH Terms). “Cervical vertebrae/diagnostic imaging” OR “Cervical” AND “spinal diseases/diagnostic imagining” (MeSH Terms) AND (chiropractic MeSH terms) or “manipulation, chiropractic”.

For CINAHL: MH “Spinal diseases/RA” AND (Cervical) OR (MH “Cervical vertebrae/RA” AND (MH “reproducibility of results” OR (MH “Reference Values”. MH “Chiropractic” OR (MH Spinal diseases/RA” AND (Cervical) OR (MH “Cervical vertebrae/RA”.

Search terms for AltHealthWatch were: (DE “Cervical vertebrae”) AND (DE “RADIOGRAPHY) OR DE “MEDICAL Radiography” and DE “REPRODUCIBLE research” OR DE “REFERENCE values”. DE “CHIROPRACTIC” OR (DE CHIROPRACTIC treatment for infant diseases” OR (DE CHIROPRACTIC treatment for juvenile diseases” OR (DE Spinal RADIOGRAPHY in chiropractic” OR (DE “SPINAL adjustment” AND (DE CERVICAL vertebrae) AND (DE “RADIOGRAPHY”) OR DE (MEDICAL radiography).

Search Terms for Web of Science were: (TS=Cervical) AND (TS=Reference values) OR (TS=reliability of results) AND (TS=X-ray) OR (TS=radiography), (TS=Cervical) AND (TS=chiropractic).

### Further Screening of Search Results, Inclusion and Exclusion Criteria

The databases searched were Pubmed (n=836), the cumulative index to nursing and allied health literature (CINAHL) (n=155), AltHealthWatch (n=0), and Web of Science (N=156).

Manual and digital deduplication assessment removed 267 articles. Articles that were found to use automation tools removed 19. Other records were removed if they were found to contain assessment of pathologies, anomalies, cancer, malignancies, bone mineral densities, and non-pertinent conditions (n=43). Following this assessment for record removal prior to screening removed 720 articles leaving 818 studies. The exclusion criteria for the remaining articles were as follows:

- Criterion 1: The study was not in English (n = 3). Note, a search was made for translations of manuscripts that were not in English and were not excluded if an English translation was found (n=0).
- Criterion 2: The study was not an *in vivo* human study (n = 4).
- Criterion 3: The study was not about radiography (n = 121).
- Criterion 4: The study was not about line drawing or mensuration for biomechanical analysis (n = 591).
- Criterion 5: The study was not on the reliability of biomechanical analysis of line drawing on lateral cervical radiographs (n = 4).

The 98 articles remaining following this search were sought for retrieval, screening, and data analysis by two independent reviewers with the third referee reviewer available to resolve discrepancies and disagreements. This retrieval found 2 duplicates and those studies were removed from further analysis. This brought the total articles which met all screening criteria for the SROL and of those remaining 96 were found by the reviewers to completely meet the study criteria and 45 were excluded. All final exclusions found were: criterion 1 (n=2), criterion 2 (n=0), criterion 3 (n=7), criterion 4 (n=13), criterion 5 (n=22). Figure 1 demonstrates the preferred reporting items for systematic review and meta-analyses (PRISMA) flow diagram for selection, inclusion and exclusion used in this SROL.

Independent reviewers DFL and JWB reviewed the remaining 96 articles according to the inclusion and exclusion criteria. Discrepancies or disagreements were resolved by the third reviewer JWH. The additional full text review found agreement on the exclusion of the 45 additional studies for criterion 1 (n=2), criterion 2 (n=0), criterion 3 (n=7), criterion 4 (n=13), and criterion 5 (n=22). The final total number of articles meeting all criteria was 51.

### Final 51 Article Characteristics

S1 Table reports the study characteristics and findings of the 51 studies meeting all inclusion and exclusion criteria. The details in the study group recorded number of investigators, techniques used for repeat analysis, methods of mensuration and parameters evaluated, and statistical results for all studies. The studies were analyzed independently by reviewers JWB and DFL and any discrepancies or disagreements were refereed by reviewer JWH.

### QAREL Instrument Analysis for Bias and Quality Assessment

The 11-item quality appraisal tool for studies of diagnostic reliability (QAREL) instrument was used to assess the studies for bias risk and quality of the studies (Table 1). Using a previously reported scoring system by Alfuth [25] and Konieczka [79]. This table has the final 51 studies of the SROL broken down by the answers to the QAREL instrument to assess quality, bias and reliability [26–76]. Each reliability study lead author and date published are listed on the left with the reference number followed by the answers to the 11 QAREL questions, the sum of the affirmative answers, the risk of bias of the study, the quality of the study, and the type of reliability type assessed within the study.

The answers to the QAREL instrument were also assessed (Figure 8, Table 2). The QAREL instrument questions most answered in the studies include: 1. Was the sample of subjects representative? (51/51 = 100.0% Yes); 2. Was the sample of raters representative? (50/51 = 98.0% Yes); 10. Was the test applied correctly and interpreted correctly? (49/51 = 96.1% Yes); and 11. Were appropriate statistical measures of agreement used? (50/51 = 98.0% Yes). The QAREL Instrument questions least answered in the studies include: 4. Were raters blinded to their own prior findings? (16/51 = 31.4% Yes); 5. Were raters blinded to the accepted reference standard? (3/51 = 5.9% Yes); 6. Were the raters blinded to the clinical information not part of the test? (12/51 = 23.5% Yes); and 7. Were raters blinded to additional non-clinical cues? (10/51 = 19.6% Yes).

**Figure 8.**
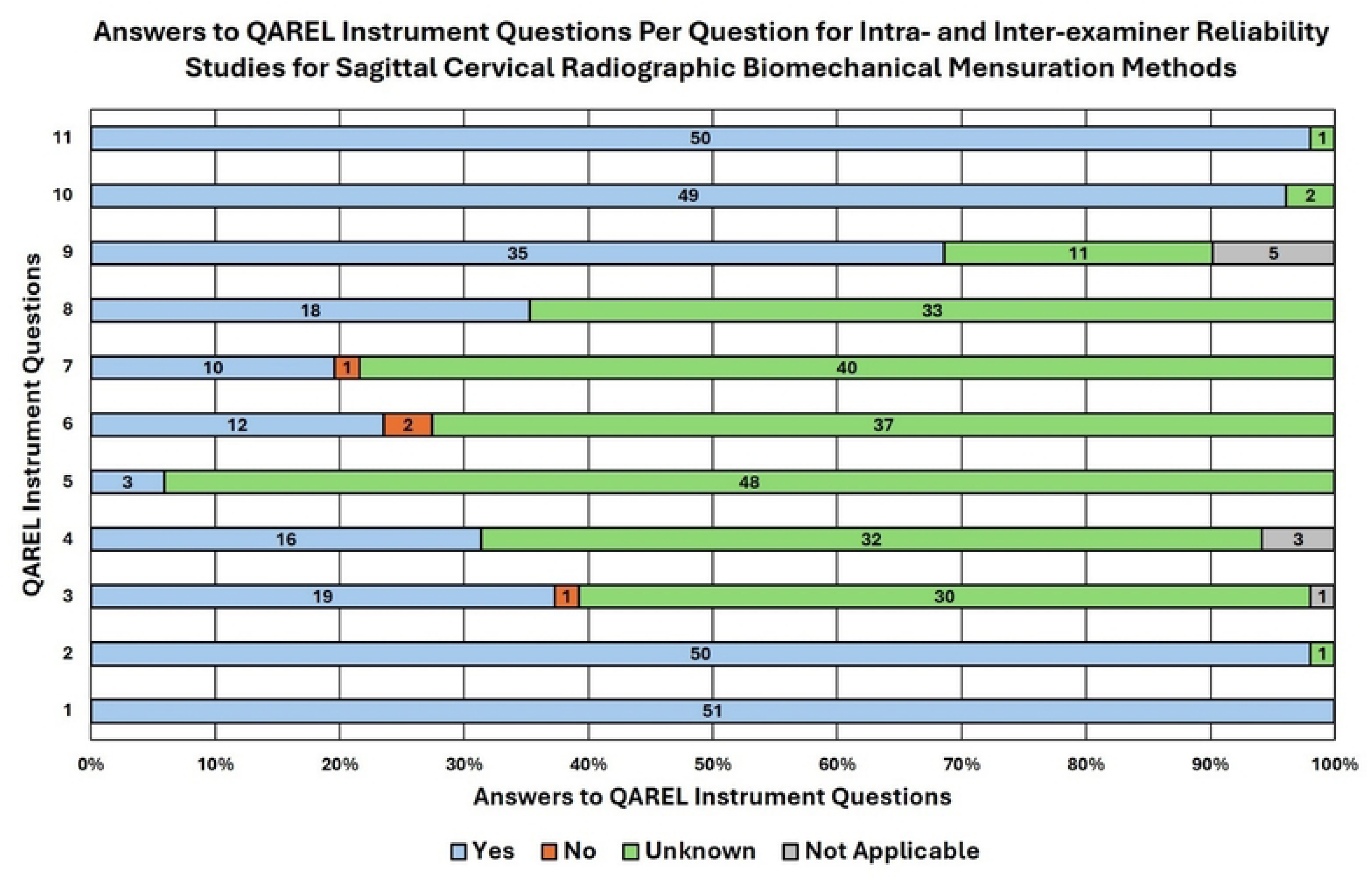
Answers to QAREL instrument questions per question for intra- and inter-examiner reliability studies for sagittal cervical radiographic biomechanical mensuration methods. This is a stacked bar chart representing the answers to QAREL instrument questions per question for intra- and inter-examiner reliability studies for sagittal cervical radiographic biomechanical mensuration methods. The stacked bars represent the percentage of studies (and show the actual number on the bar) that provided the respective answer (x-axis; see legend for color) per QAREL instrument question (y-axis).

**Table 2.** Answers to QAREL instrument questions per question for intra- and inter-examiner reliability studies for sagittal cervical radiographic biomechanical mensuration methods and study quality and reliability type.

| QAREL Answers |  | QAREL Instrument Questions |  |  |  |  |  |  |  |  |  |  | Study Quality |  | Study Reliability |  |
| --- | --- | --- | --- | --- | --- | --- | --- | --- | --- | --- | --- | --- | --- | --- | --- | --- |
|  |  | 1 | 2 | 3 | 4 | 5 | 6 | 7 | 8 | 9 | 10 | 11 |  |  |  |  |
| Yes |  | 51 | 50 | 19 | 16 | 3 | 12 | 10 | 18 | 35 | 49 | 50 | L | 9 | IA | 6 |
| No |  | 0 | 0 | 1 | 0 | 0 | 2 | 1 | 0 | 0 | 0 | 0 | M | 27 | IR | 10 |
| Unknown |  | 0 | 1 | 30 | 32 | 48 | 37 | 40 | 33 | 11 | 2 | 1 | H | 15 | B | 35 |
| Not Applicable |  | 0 | 0 | 1 | 3 | 0 | 0 | 0 | 0 | 5 | 0 | 0 |  |  |  |  |

L = Low; M = Moderate; H = High; IA = Intra-examiner; IR = Inter-examiner; B = Both (Intra- and Inter-examiner)

When assessing intra- and inter-examiner reliability (n=51) articles included in the SROL, it was found that 15 (29.4%) articles had a low risk of bias assessment (high quality), 27 (52.9%) articles were found to have a moderate risk of bias following assessment (moderate quality), and 9 (17.6%) studies were found to have a high-risk bias assessed (low quality) (Figure 9).

**Figure 9.**
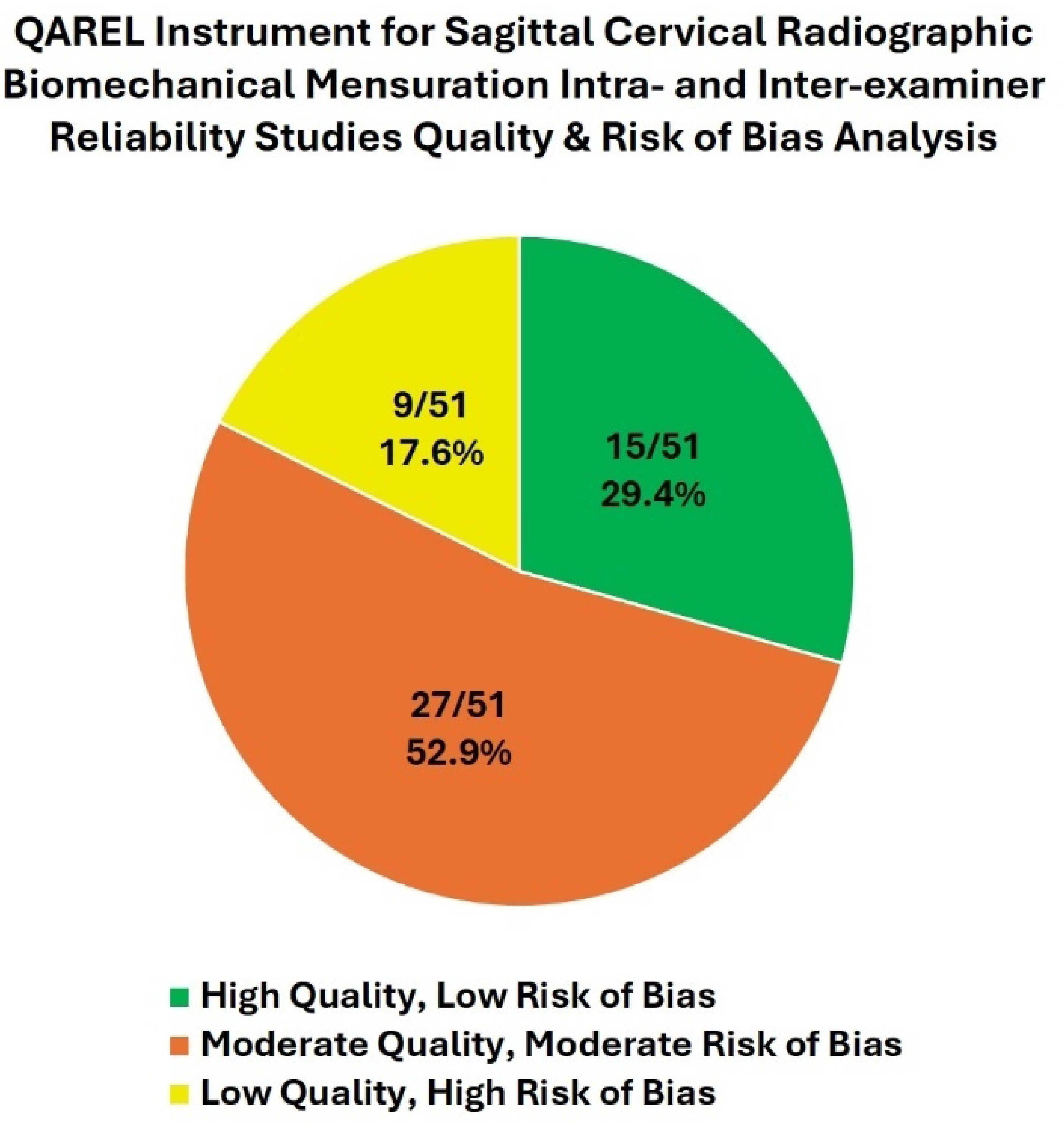
Sagittal cervical radiographic biomechanical mensuration intra- and inter-examiner reliability studies quality and risk of bias analysis using the QAREL instrument.

When assessing intra-examiner reliability (n=41) articles included in the SROL, it was found that 13 (31.7%) articles had a low risk of bias assessment (high quality), 22 (53.7%) articles were found to have a moderate risk of bias following assessment (moderate quality), and 6 (14.6%) studies were found to have a high-risk bias assessed (low quality) (Figure 10).

**Figure 10.**
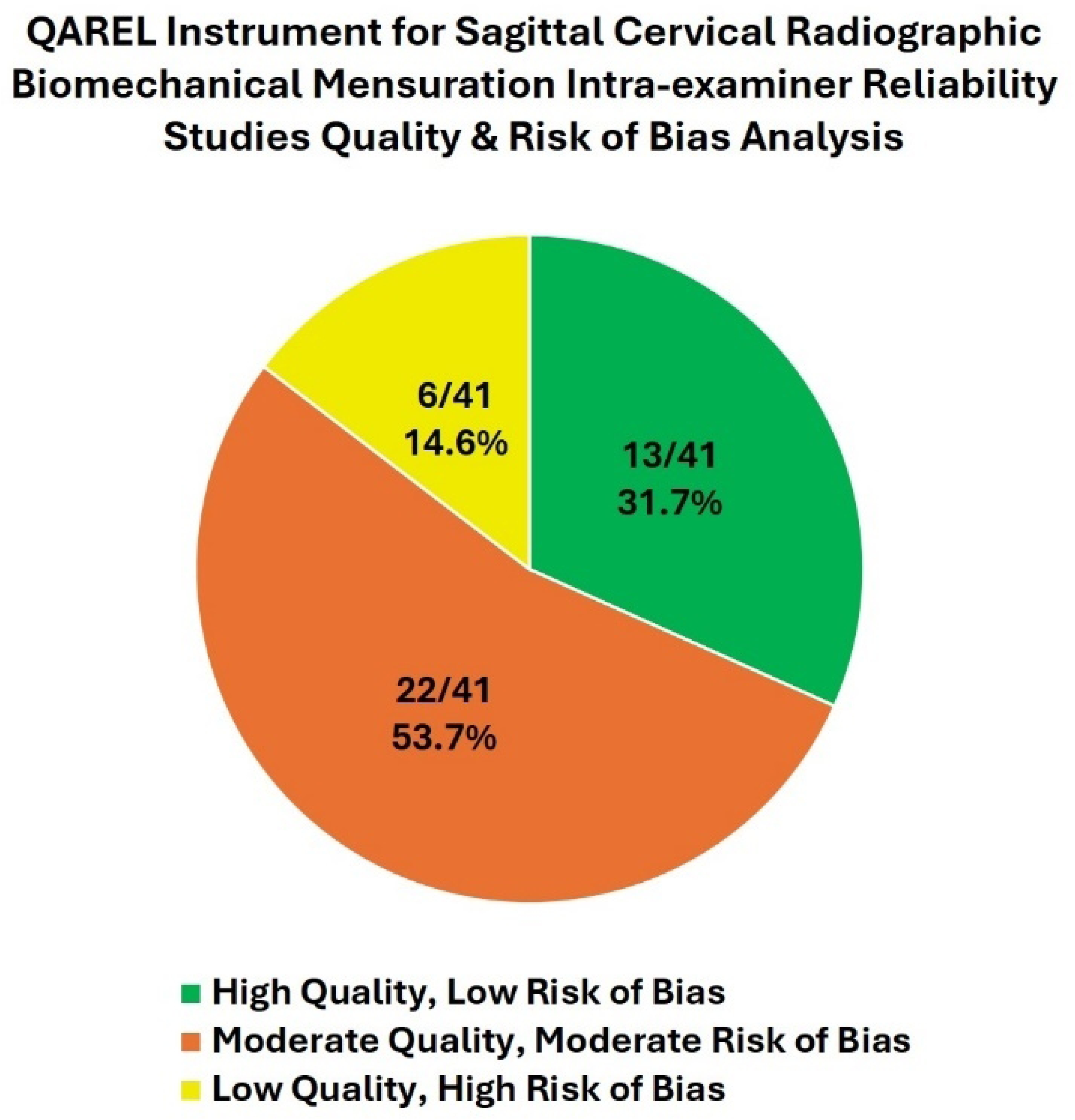
Sagittal cervical radiographic biomechanical mensuration intra-examiner reliability studies quality and risk of bias analysis using the QAREL instrument.

When assessing inter-examiner reliability (n=45) articles included in the SROL, it was found that 14 (31.1%) articles had a low risk of bias assessment (high quality), 23 (51.1%) articles were found to have a moderate risk of bias following assessment (moderate quality), and 8 (17.8%) studies were found to have a high-risk bias assessed (low quality) (Figure 11).

**Figure 11.**
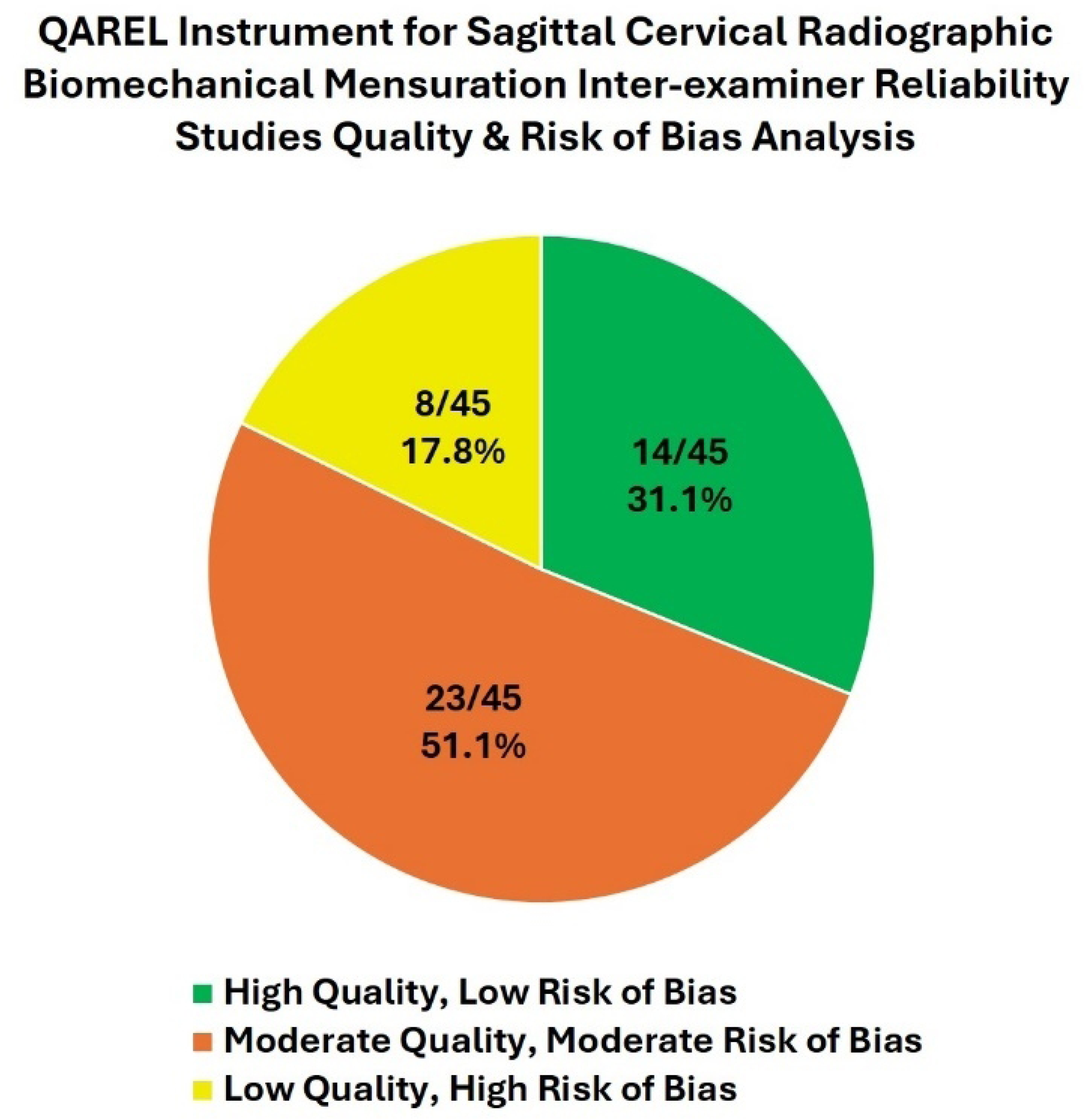
Sagittal cervical radiographic biomechanical mensuration inter-examiner reliability studies quality and risk of bias analysis using the QAREL instrument.

Intra-examiner reliability studies using Cobb method C2-C7 reported 4 good and 7 excellent Intraclass Correlation Coefficients (ICC). Intra-examiner reliability studies using Cobb method C1-C7 reported 3 excellent ICCs. Intra-examiner reliability studies using Posterior Tangent (C2-C7) method reported 8 excellent ICCs. Intra-examiner reliability studies using C0-C2 reported 1 moderate and 2 excellent ICCs. Intra-examiner reliability studies using Sagittal Vertical Axis (C2-C7) reported 3 excellent ICCs. Intra-examiner reliability studies using T1 Slope reported 1 moderate, 1 good, and 4 excellent ICCs. Intra-examiner reliability studies using Thoracic Inlet reported 2 good and 2 excellent ICCs. Intra-examiner reliability studies using Intersegmental Translation reported 1 good ICC. Intra-examiner reliability studies using Intersegmental Rotation studies reported 6 good and 1 excellent ICCs (Figure 6, S2 Table).

Inter-examiner reliability studies using Cobb method C2-C7 reported 7 good and 6 excellent Intraclass Correlation Coefficients (ICC). Inter-examiner reliability studies using Cobb method C1-C7 reported 4 excellent ICCs. Inter-examiner reliability studies using Posterior Tangent (C2-C7) method reported 2 good and 5 excellent ICCs. Inter-examiner reliability studies using C0-C2 reported 1 moderate, 2 good, and 1 excellent ICCs. Inter-examiner reliability studies using Sagittal Vertical Axis (C2-C7) reported 5 excellent ICCs. Inter-examiner reliability studies using T1 Slope reported 3 good and 3 excellent ICCs. Inter-examiner reliability studies using Thoracic Inlet reported 3 good and 1 excellent ICCs. Inter-examiner reliability studies using Intersegmental Translation reported 2 good ICC. Inter-examiner reliability studies using Intersegmental Rotation studies reported 6 good ICCs (Figure 4, S3 Table).

### Intra-and Inter-examiner Reliability with SEM and MAD

Assessment of intra-examiner reliability studies for standard error of measurement (SEM) and mean absolute distance (MAD) found the studies of moderate to high quality reported ranges of SEM in angular measurements from 0.5-5.47° and MAD in angular measurements from 0.35-3.8° and translation measurements from 0.175-0.3 mm (Figure 7, S2 Table).

The biomechanical mensuration method that showed the lowest SEM of angular measurements was Posterior Tangent (C2-C7) method with 0.50°. There were no SEMs reported of translational measurements. The biomechanical mensuration method that showed the highest SEM of angular measurements was Cobb Method (C2-C7) with 5.47°. The biomechanical mensuration method that showed the lowest MAD of angular measurements was Intersegmental Rotation with 0.35°. The biomechanical mensuration method that showed the lowest MAD of translation measurements was Intersegmental Translation with 0.175 mm.

The biomechanical mensuration method that showed the highest SEM of angular measurements was Cobb method (C2-C7) with 3.8°. There were no SEMs reported of translational measurements. The biomechanical mensuration method that showed the highest MAD of angular measurements was Cobb Method (C2-C7) with 3.8°. The biomechanical mensuration method that showed the highest MAD of translation measurements was sagittal vertical axis (C2-C7) with 0.3 mm.

The biomechanical mensuration method that showed the greatest precision of SEMs across all studies in angular measurements was intersegmental rotation with SEMs ranging from 0.79-1.635°. The Cobb method (C2-C7) was the only angular measurement method with more than 1 study with MADs reported and ranged from 0.8-3.8°. All translation measurement methods with MADs only had 1 study. The biomechanical mensuration method that showed the least precision of SEMs across all studies in angular measurements was Cobb method (C2-C7) with SEMs ranging from 2.06-5.47°.

Assessment of inter-examiner reliability studies for mean absolute distance (MAD) and standard error of measurement (SEM) found the studies of moderate to high quality reported ranges of SEM in angular measurements from 0.79-7.0° and MAD in angular measurements from 0.455-5.2° and translation measurements of 0.345 mm (Figure 5, S3 Table).

The biomechanical mensuration method that showed the lowest SEM of angular measurements was intersegmental rotation with 0.79°. There were no SEMs reported of translational measurements. The biomechanical mensuration method that showed the highest SEM of angular measurements was Cobb method (C2-C7) with 7.0°. The biomechanical mensuration method that showed the lowest MAD of angular measurements was intersegmental rotation with 0.35°. Intersegmental translation was the only study that reported a MAD of translation measurements with 0.345 mm.

The biomechanical mensuration method that showed the highest SEM of angular measurements was Cobb method (C2-C7) with 7.0°. There were no SEMs reported of translational measurements. The biomechanical mensuration method that showed the highest MAD of angular measurements was Cobb method (C2-C7) with 5.2°.

The biomechanical mensuration method that showed the greatest precision of SEMs across all studies in angular measurements was intersegmental rotation with SEMs ranging from 0.79-1.805°. The Cobb method (C1-C7) was the only angular measurement method with more than 1 study with MADs reported and ranged from 0.9-3.4°. The biomechanical mensuration method that showed the least precision of SEMs across all studies in angular measurements was Cobb Method (C2-C7) with SEMs ranging from 2.71-7.0°.

### Variables of Importance

A systematic review of the literature discovered 1147 articles related to lateral cervical spine biomechanical mensuration. Following de-duplication, inclusion and exclusion analysis, 51 articles were found to meet all criteria. The use of the QAREL tool distinguished articles for quality and bias. Two initial reviewers and a third referee reviewer assessed the remaining studies to determine the reliability of several commonly used radiographic measurements of the cervical spine. Good-to-excellent reliability was found in the lateral cervical spine mensuration studies. Regulatory boards and healthcare practitioners, physicians, and clinicians who treat cervical spine disorders can depend on biomechanical mensuration methods of sagittal cervical radiography to exhibit good-to-excellent reliability within and between examiners.

## Discussion

Our hypothesis was that the available literature reporting on the on the intra- and inter-examiner reliability of the biomechanical assessment of the sagittal cervical spine on radiography used in clinical practice provides sufficient low bias risk, high-quality studies that establish good or better reliability. The results of this SROL confirm this hypothesis and the null hypothesis is rejected. Prior rapid reviews, narrative reviews, and selective review studies show erroneous conclusions regarding the reliability and validity of radiographic mensuration [84–88]. The biomechanical analysis of the sagittal cervical, cranio-cervical, and cervico-thoracic spine are important clinical findings included in the triage, diagnosis, and management (treatment, referrals, etc.) of cervical spine disorders. Further, manual, computer-assisted, and machine learning studies confirm good-to-excellent reliability of cervical spine mensuration methods [89,90].

### Conglomeration and Heterogeneity of Reliability Measure Findings

The explanation of heterogeneity of findings is important in this SROL. Current studies are adapting the use of machine learning and digital analysis for confirmation of reliability of cervical spine mensuration. Studies in this SROL incorporated both manual and digital measured assessments. There can be significant differences ICCs, between manually assessed spine parameters and computer-aided mensuration methods. Although previous studies have shown that manual methods provide lower reliability, there is excellent reliability with computer-aided mensuration, including studies with R^2^=1 (perfect correlation) models for machine learning programs [90]. Other sources of heterogeneity can often be explained by the quality and type of image used, whether digital or pain film radiographs. The source of the image can alter mensuration including x-ray radiographs versus EOS low dose images or biplanar stereoradiography. Heterogeneity of findings can also be explained by differences in the statistical analysis and reportage. We included studies that use ICCs or Pearson correlation coefficients and other studies that used Cohen’s kappa agreement or Bland-Altman plots; all of which can have different reliability assessment and interpretation [77].

### Systematic Review Significance and Clinical Application

This SROL discovered good-to-excellent intra- and inter-examiner reliability of biomechanical mensuration methods using cervical spine radiography. It seems this SROL is the first in the body of knowledge to provide this complete, comprehensive analysis. The use of the preferred reporting items for systematic reviews and meta-analyses (PRISMA) checklist for this SROL improves quality, reduces erroneous conclusions, and improves confidence in assessment [86]. The use of sagittal cervical radiography is widespread for clinicians and physicians who treat cervical spine disorders. Sagittal cervical biomechanical measurements are used by entry level and trauma clinicians [91], primary care providers [92], surgeons [93–95], physical therapists [96,97], chiropractors [97], device manufacturers [98], modeling and machine learning science [89,90,93,97], third-party payors [98,99], governmental bodies [100], and educational institutions [99].

Due to the significant contribution of neck pain to GBD and YLD, reliability of diagnostic assessments is essential. Unhealthy, abnormal sagittal cervical spine biomechanics and measurements can cause and are associated with altered health quality of life, pain, and dysfunction and disability. Recent studies on abnormal forward head posture (FHP), loss of cervical lordosis, and abnormal spine alignment have shown association with numerous disorders and abnormal health outcomes. Abnormal FHP has been associated with altered resting brain state [101], reduced overall human performance and mental activity [101,102], neck pain [103], altered gait and postural control [104], decrease cerebral blood perfusion and hemodynamic control [104,105], postural dislocation [107], temporomandibular joint disorders [108,109], and respiratory function [109].

Physicians providing interventions to improve sagittal cervical alignment and posture rely on cervical spine radiography for the triage, diagnosis, management (treatment and referrals), prognosis, and progress evaluation of cervical spine disorders [94]. The application of these techniques requires lateral cervical radiographic mensuration to determine specific treatment strategies and to monitor progress [110–114]. Studies have indicated the importance of cervical spine and thoracic inlet morphology to assess the relative ideal sagittal cervical spinal alignment and posture for a given patient which can vary [115]. The morphological assessment of the cervical spine requires reliable radiographic mensuration of sagittal cervical spine parameters [115,116]. Recent advancements in artificial intelligence (AI) and machine learning technology have also improved the speed and reliability of biomechanical mensuration of the cervical spine [90]. It is expected that future advances in this technology will continue to assist decision-making and improve clinical certainty in the triage, diagnosis, management (treatment and referrals), prognosis, and progress evaluation of cervical spine disorders [117–119].

### Previous “Reviews of Literature” Are Flawed and Report Erroneous Conclusions

Imaging studies have been used as a high standard in the diagnosis and treatment of cervical spine disorders [120,121]. This current SROL improves the understanding of the reliability of cervical spine biomechanical mensuration and is in contrast to prior studies that have wrongly asserted that x-ray line drawing has no clear reliability [84–88]. The use of sagittal cervical radiography to produce diagnostic images can be encouraged as the results of the biomechanical mensuration will improve patient diagnosis, treatment options and outcomes [121]. Despite this clinical necessity for the use of spine radiography, there have been some recent studies that have made erroneous statements about the reliability, validity, and use of radiography for neck pain and other spinal musculoskeletal conditions [7,84–88,122]. For example, regarding radiographic measurements of cervical lordosis (Fig 2) that are reported in the scientific literature there is often conflicting validity evidence reported in the literature [122,123].

In their 2018 systematic review with meta-analysis, Guo et al. [122] reported that there is no difference in radiographically measured cervical lordotic values between patients with chronic neck pain compared to asymptomatic controls. Problematically, Guo et al. [122] inappropriately cited and included the Harrison et al. 1996 [124] investigation as a case control when in fact this manuscript did not report such data and instead, they [122] failed to properly identify the Harrison et al 2004 [125] investigation as the proper case control to be included in their [112] analysis. This gross oversight led to their [112] erroneous conclusion that radiographic cervical lordosis is not a significant predictor of neck pain. In contrast, the 2024 systematic review and meta-analysis by Kim et al. [123] incorporated the 2004 Harrison et al. [125] case control investigation in their analysis and concluded that patients with neck pain have a statistically and clinically significant reduced cervical lordosis on X-ray. Thus, while there exists evidence to the contrary, the current authors argue that the prevailing literature continues to support the use of sagittal spine radiography to analyze, diagnose, and treat patients with cervical spine disorders [123,125–128]. The results of our current SROL establishes the reliability of the various x-ray measurements for sagittal cervical spine displacements and provides the foundation for continued investigations into the validity of cervical deformity evaluations from a displacement perspective.

### Strength, Limitations, and Future Research Recommendations

This SROL followed the PRISMA study design and checklists for reporting of systematic reviews and meta-analyses. This SROL was registered with the international prospective register of systematic reviews (PROSPERO) prior to any article search. This SROL performed and extensive literature search from numerous health and biotechnology article databases. The thorough and multi-layered search strategy yielded 51 articles meeting the inclusion and exclusion criteria. This large number of studies is rare for most SROLs. Additional strength of the SROL is the use of the synthesis without meta-analysis (SWiM) guidelines for presentation of subgroup assessments for intra- and inter-examiner reliability. Additionally, the use of two separate reviewers and a third referee reviewer for consensus was a strong feature of our design. The use of the QAREL tool for risk of bias and quality assessment makes the SROL more robust and the conclusions more accurate. Future SROLs should include a similar multi-layer methodology to ensure quality of conclusions.

This SROL addresses the elements of PRISMA checklists and encompasses a robust study count of 51 on the reliability of biomechanical mensuration of the sagittal cervical spine on radiography used in clinical practice. That said, there are several biomechanical mensuration methods included which diminishes the number per method. Many studies did not document or calculate SEM and MAD for their methods of measurement and the included studies used different statistical analyses to establish reliability. Also, given the absence of meta-analyses on the topic of reliability of radiographic mensuration, the investigators were not able to perform a grey analysis of possible crossover between upright standing lateral cervical radiographs and other cervical spine imaging. That said, reliability was nonetheless evaluated in the non-crossover studies [129]. No formal meta-analysis was performed in this study either which would be a good follow-up to perform from the included studies of this SROL. Other limitations to be considered are those in the studies that meet the inclusion criteria. Reliability studies need to make sure they have calculated a power analysis to ensure sufficient power of their results and that they are answering all questions of the QAREL checklist (or any other reliability study checklist) to ensure the highest of quality and lowest risk of bias (Fig 1, Table 2). Not all studies included met these criteria. This analysis provides a clearly delineated guide to rate the quality of an article and should be applied to future research.

Future reliability studies on biomechanical mensuration methods need to include SEM and MAD for proper study results and to facilitate further meta-analysis. Researchers should continue to perform SROLs and meta-analyses on the reliability of biomechanical mensuration methods in all global and regional sagittal and anteroposterior spinal imaging. An essential element of validity of measurements is that the measurement methods are reliable.

### Conclusion

The methodology and design features found the preponderance of evidence for intra- and inter-examiner reliability for the biomechanical mensuration of the human sagittal cervical spine alignment to have good-to excellent results. The robust nature of our findings demonstrate that the use of sagittal cervical spine radiography is a reliable tool to aid in the diagnosis and treatment recommendations for cervical spine and associated disorders.

## Data Availability

Data is available upon reasonable request from the corresponding author.

## Acknowledgments

This research was funded by CBP NonProfit and partially funded by a Grant from Australian Spinal Research Foundation.

## Supporting Information

**S1 Table. Study Characteristics and Findings of the Reliability Studies on Biomechanical Mensuration Methods of Sagittal Cervical Radiography Used in Clinical Practice**

**S2 Table. Sub-Group Analysis of Intra-examiner Reliability Studies on Biomechanical Mensuration Methods of Sagittal Cervical Radiography by Intra-examiner Reliability Statistics, Study Quality, SEM, and MAD**

**S3 Table. Sub-Group Analysis of Inter-examiner Reliability Studies on Biomechanical Mensuration Methods of Sagittal Cervical Radiography by Inter-examiner Reliability Statistics, Study Quality, SEM, and MAD**

## References

1. Shin DW, Shin JI, Koyanagi A, Jacob L, Smith L, Lee H, et al. Global, regional, and national neck pain burden in the general population, 1990-2019: An analysis of the global burden of disease study 2019. Front Neurol. 2022;13:955367. doi:10.3389/fneur.2022.955367

2. Kazeminasab S, Nejadghaderi SA, Amiri P, Pourfathi H, Araj-Khodaei M, Sullman MJM, et al. Neck pain: global epidemiology, trends and risk factors. BMC Musculoskelet Disord. 2022;23(1):26. doi:10.1186/s12891-021-04957-4

3. GBD 2021 Neck Pain Collaborators. Global, regional, and national burden of neck pain, 1990-2020, and projections to 2050: a systematic analysis of the Global Burden of Disease Study 2021. Lancet Rheumatol. 2024;6(3):e142-e155. doi:10.1016/S2665-9913(23)00321-1

4. Igwesi-Chidobe CN, Effiong E, Umunnah JO, Ozumba BC. Occupational biopsychosocial factors associated with neck pain intensity, neck-disability, and sick leave: A cross-sectional study of construction labourers in an African population. PLoS One. 2024;19(4):e0295352. doi:10.1371/journal.pone.0295352

5. Harrison DE, Oakley PA, Moustafa IM. Don’t throw the ‘Bio’ out of the Bio-Psycho-Social model: Editorial for spine rehabilitation in 2022 and beyond. J Clin Med. 2023;12(17):5602. doi:10.3390/jcm12175602

6. Luque-Suarez A, Falla D, Morales-Asencio JM, Martinez-Calderon J. Is kinesiophobia and pain catastrophising at baseline associated with chronic pain and disability in whiplash-associated disorders? A systematic review. Br J Sports Med. 2020;54(15):892–897. doi:10.1136/bjsports-2018-099569

7. Chou R, Côté P, Randhawa K, Torres P, Yu H, Nordin M, et al. The Global Spine Care Initiative: applying evidence-based guidelines on the non-invasive management of back and neck pain to low-and middle-income communities. Eur Spine J. 2018;27(Suppl 6):851–860. doi:10.1007/s00586-017-5433-8

8. Oxland TR. Fundamental biomechanics of the spine--What we have learned in the past 25 years and future directions. J Biomech. 2016;49(6):817–832. doi:10.1016/j.jbiomech.2015.10.035

9. Hesselink JR. Spine imaging: history, achievements, remaining frontiers. AJR Am J Roentgenol. 1988;150(6):1223–1229. doi:10.2214/ajr.150.6.1223

10. Pellisé F, Vila-Casademunt A, Ferrer M, Domingo-Sàbat M, Bagó J, Pérez-Grueso FJ, et al. Impact on health related quality of life of adult spinal deformity (ASD) compared with other chronic conditions. Eur Spine J. 2015;24:3–11. doi:10.1007/s00586-014-3542-1

11. Kyrölä K, Repo J, Mecklin JP, Ylinen J, Kautiainen H, Häkkinen A. Spinopelvic changes based on the simplified SRS-Schwab Adult Spinal Deformity Classification: relationships with disability and health-related quality of life in adult patients with prolonged degenerative spinal disorders. Spine (Phila Pa 1976). 2018;43(7):497-502. doi:10.1097/BRS.0000000000002370

12. Protopsaltis TS, Scheer JK, Terran JS, Smith JS, Hamilton DK, Kim HJ, et al. How the neck affects the back: Changes in regional cervical sagittal alignment correlate to HRQOL improvement in adult thoracolumbar deformity patients at 2-year follow-up. J Neurosurg Spine. 2015;23(2):153–8. doi:10.3171/2014.11.SPINE1441.

13. Ling FP, Chevillotte T, Leglise A, Thompson W, Bouthors C, Le Huec JC. Which parameters are relevant in sagittal balance analysis of the cervical spine? A literature review. Eur Spine J. 2018;27(Suppl 1):8–15. doi:10.1007/s00586-018-5462-y.

14. Moustafa IM, Shousha T, Arumugam A, Harrison DE. Is thoracic kyphosis relevant to pain, autonomic nervous system function, disability, and cervical sensorimotor control in patients with chronic nonspecific neck pain? J Clin Med. 2023;12(11):3707. doi:10.3390/jcm12113707.

15. Kaale BR, McArthur TJ, Barbosa MH, Freeman MD. Post-traumatic atlanto-axial instability: A combined clinical and radiological approach for the diagnosis of pathological rotational movement in the upper cervical spine. J Clin Med. 2023;12(4):1469. doi:10.3390/jcm12041469.

16. Moustafa IM, Diab AAM, Harrison DE. Does improvement towards a normal cervical sagittal configuration aid in the management of lumbosacral radiculopathy: A randomized controlled trial. J Clin Med. 2022;11(19):5768. doi:10.3390/jcm11195768.

17. Arnone PA, Kraus SJ, Farmen D, Lightstone DF, Jaeger J, Theodossis C. Examining clinical opinion and experience regarding utilization of plain radiography of the spine: evidence from surveying the chiropractic profession. J Clin Med. 2023;12(6):2169. doi:10.3390/jcm12062169.

18. Oakley PA, Moustafa IM, Harrison DE. The influence of sagittal plane spine alignment on neurophysiology and sensorimotor control measures: optimization of function through structural correction. In: Bernardo-Filho M, editor. Therapy Approaches in Neurological Disorders. London: IntechOpen; 2021.

19. Kuo DT, Tadi P. Cervical spondylosis. [Updated 2023 May 1]. In: StatPearls [Internet]. Treasure Island (FL): StatPearls Publishing; 2024 Jan–. Available from: https://www.ncbi.nlm.nih.gov/books/NBK551557/

20. Torlincasi AM, Waseem M. Cervical injury. [Updated 2022 Aug 22]. In: StatPearls [Internet]. Treasure Island (FL): StatPearls Publishing; 2024 Jan–. Available from: https://www.ncbi.nlm.nih.gov/books/NBK448146/

21. Kim HJ, Virk S, Elysee J, Ames C, Passias P, Shaffrey C, et al. Surgical strategy for the management of cervical deformity is based on type of cervical deformity. J Clin Med. 2021;10(21):4826. doi:10.3390/jcm10214826.

22. Foley D, Hardacker P, McCarthy M. Emerging technologies within spine surgery. Life (Basel). 2023;13(10):2028. doi:10.3390/life13102028.

23. Liu C, Zhu W, Li Y, Li X, Shi B, Kong C, et al. How does cervical sagittal profile change after the spontaneous compensation of global sagittal imbalance following one-or two-level lumbar fusion. BMC Musculoskelet Disord. 2024;25(1):387. doi:10.1186/s12891-024-07518-7.

24. Page MJ, McKenzie JE, Bossuyt PM, Boutron I, Hoffmann TC, Mulrow CD, et al. The PRISMA 2020 statement: an updated guideline for reporting systematic reviews. Syst Rev. 2021;10(1):89. doi:10.1186/s13643-021-01626-4.

25. McGowan J, Sampson M, Salzwedel DM, Cogo E, Foerster V, Lefebvre C. PRESS peer review of electronic search strategies: 2015 guideline statement. J Clin Epidemiol. 2016;75:40–6. doi:10.1016/j.jclinepi.2016.01.021.

26. Alhilali LM, Fakhran S. Evaluation of the intervertebral disk angle for the assessment of anterior cervical diskoligamentous injury. AJNR Am J Neuroradiol. 2013;34(12):2399–404. doi:10.3174/ajnr.A3585.

27. Andriola FO, Kulczynski FZ, Deon PH, Melo DADS, Zanettini LMS, Pagnoncelli RM. Changes in cervical lordosis after orthognathic surgery in skeletal class III patients. J Craniofac Surg. 2018;29(6):e598–603. doi:10.1097/SCS.0000000000004644.

28. Balouch E, Burapachaisri A, Woo D, Norris Z, Segar A, Ayres EW, et al. Assessing postoperative pseudarthrosis in anterior cervical discectomy and fusion (ACDF) on dynamic radiographs using novel angular measurements. Spine (Phila Pa 1976). 2022;47(16):1151-6. doi:10.1097/BRS.0000000000004375.

29. Barth M, Weiß C, Brenke C, Schmieder K. Reliability and scientific use of a surgical planning software for anterior cervical discectomy and fusion (ACDF). Eur Spine J. 2017;26(4):1305–11. doi:10.1007/s00586-017-4957-2.

30. Bellabarba C, Karim F, Tavolaro C, Zhou H, Bremjit P, Nguyen QT, et al. The mandible-C2 angle: a new radiographic assessment of occipitocervical alignment. Spine J. 2021;21(1):105–13. doi:10.1016/j.spinee.2020.07.003.

31. Bono CM, Schoenfeld A, Rampersaud R, Levi A, Grauer J, Arnold P, et al. Reproducibility of radiographic measurements for subaxial cervical spine trauma. Spine (Phila Pa 1976). 2011;36(17):1374-9. doi:10.1097/BRS.0b013e318221e169.

32. Cannada LK, Scherping SC, Yoo JU, Jones PK, Emery SE. Pseudoarthrosis of the cervical spine: a comparison of radiographic diagnostic measures. Spine (Phila Pa 1976). 2003;28(1):46-51. doi:10.1097/00007632-200301010-00012.

33. Chamnan R, Chantarasirirat K, Paholpak P, Wiley K, Buser Z, Wang JC. Occipitocervical measurements: correlation and consistency between multi-positional magnetic resonance imaging and dynamic radiographs. Eur Spine J. 2020;29(11):2795–803. doi:10.1007/s00586-020-06415-6.

34. Champain S, Benchikh K, Nogier A, Mazel C, Guise JD, Skalli W. Validation of new clinical quantitative analysis software applicable in spine orthopaedic studies. Eur Spine J. 2006;15(6):982–91. doi:10.1007/s00586-005-0927-1.

35. Chen L, Lan Z, Xu X, Lin J, Hu H. Accuracy and repeatability of computer aided cervical vertebra landmarking in cephalogram. J Huazhong Univ Sci Technolog Med Sci. 2012;32(1):119–23. doi:10.1007/s11596-012-0021-y.

36. Côté P, Cassidy JD, Yong-Hing K, Sibley J, Loewy J. Apophysial joint degeneration, disc degeneration, and sagittal curve of the cervical spine. Can they be measured reliably on radiographs? Spine (Phila Pa 1976). 1997;22(8):859–64. doi:10.1097/00007632-199704150-00007.

37. Daffin L, Stuelcken M, Sayers M. Internal and external sagittal craniovertebral alignment: A comparison between radiological and photogrammetric approaches in asymptomatic participants. Musculoskelet Sci Pract. 2019;43:12–7. doi:10.1016/j.msksp.2019.05.003.

38. Donk RD, Fehlings MG, Verhagen WIM, Arnts H, Groenewoud H, Verbeek ALM, et al. An assessment of the most reliable method to estimate the sagittal alignment of the cervical spine: analysis of a prospective cohort of 138 cases. J Neurosurg Spine. 2017;26(5):572–6. doi:10.3171/2016.10.SPINE16632.

39. Douglas TS, Sanders V, Machers S, Pitcher R, van As AB. Digital radiographic measurement of the atlantodental interval in children. J Pediatr Orthop. 2007;27(1):23–6. doi:10.1097/01.bpo.0000242443.42176.67.

40. Dvorak J, Froehlich D, Penning L, Baumgartner H, Panjabi MM. Functional radiographic diagnosis of the cervical spine: flexion/extension. Spine (Phila Pa 1976). 1988;13(7):748–55. doi:10.1097/00007632-198807000-00007.

41. Frobin W, Leivseth G, Biggemann M, Brinckmann P. Sagittal plane segmental motion of the cervical spine. A new precision measurement protocol and normal motion data of healthy adults. Clin Biomech (Bristol, Avon). 2002;17(1):21–31. doi:10.1016/s0268-0033(01)00105-x.

42. Gadotti IC, Armijo-Olivo S, Silveira A, Magee D. Reliability of the craniocervical posture assessment: visual and angular measurements using photographs and radiographs. J Manipulative Physiol Ther. 2013;36(9):619–25. doi:10.1016/j.jmpt.2013.09.002.

43. Gadotti IC, Magee D. Assessment of intrasubject reliability of radiographic craniocervical posture of asymptomatic female subjects. J Manipulative Physiol Ther. 2013;36(1):27–32. doi:10.1016/j.jmpt.2012.12.009.

44. Guyard T, Le Quellec A, Garrigues F, Saraux A. Reproducibility and diagnostic value of a new method using ratios to diagnose anterior atlanto-axial subluxation on plain radiographs. Joint Bone Spine. 2021;88(5):105229. doi:10.1016/j.jbspin.2021.105229.

45. Gwinn DE, Iannotti CA, Benzel EC, Steinmetz MP. Effective lordosis: analysis of sagittal spinal canal alignment in cervical spondylotic myelopathy. J Neurosurg Spine. 2009;11(6):667–72. doi:10.3171/2009.7.SPINE08656.

46. Hardacker JW, Shuford RF, Capicotto PN, Pryor PW. Radiographic standing cervical segmental alignment in adult volunteers without neck symptoms. Spine (Phila Pa 1976). 1997;22(13):1472–80; discussion 1480. doi:10.1097/00007632-199707010-00009.

47. Harrison DE, Harrison DD, Cailliet R, Troyanovich SJ, Janik TJ, Holland B. Cobb method or Harrison posterior tangent method: which to choose for lateral cervical radiographic analysis. Spine (Phila Pa 1976). 2000;25(16):2072–8. doi:10.1097/00007632-200008150-00011.

48. Herman S. Computed tomography contrast enhancement principles and the use of high-concentration contrast media. J Comput Assist Tomogr. 2004;28 Suppl 1:S7–11. doi:10.1097/01.rct.0000120855.80935.2f.

49. Jackson BL, Harrison DD, Robertson GA, Barker WF. Chiropractic biophysics lateral cervical film analysis reliability. J Manipulative Physiol Ther. 1993;16(6):384–91.

50. Janusz P, Tyrakowski M, Yu H, Siemionow K. Reliability of cervical lordosis measurement techniques on long-cassette radiographs. Eur Spine J. 2016;25(11):3596–601. doi:10.1007/s00586-015-4345-8.

51. Jiang SD, Chen JW, Yang YH, Chen XD, Jiang LS. Intraobserver and interobserver reliability of measures of cervical sagittal rotation. BMC Musculoskelet Disord. 2014;15:332. doi:10.1186/1471-2474-15-332.

52. Johnson GM. The correlation between surface measurement of head and neck posture and the anatomic position of the upper cervical vertebrae. Spine (Phila Pa 1976). 1998;23(8):921–7. doi:10.1097/00007632-199804150-00015.

53. Kuhr M, Hohmann D, Schramm M, Martus P. Radiographic evaluation of the upper cervical spine in rheumatoid arthritis: a retrospective analysis. Eur Spine J. 1996;5(2):107–11. doi:10.1007/BF00298389.

54. Kunakornsawat S, Pluemvitayaporn T, Pruttikul P, Punpichet S, Piyasakulkaew C, Arirachakaran A, et al. A new method for measurement of occipitocervical angle by occiput-C3 angle. Eur J Orthop Surg Traumatol. 2017;27(8):1051–6. doi:10.1007/s00590-016-1881-9.

55. Lafage R, Ferrero E, Henry JK, Challier V, Diebo B, Liabaud B, et al. Validation of a new computer-assisted tool to measure spino-pelvic parameters. Spine J. 2015;15(12):2493–502. doi:10.1016/j.spinee.2015.08.067.

56. Lee HD, Jeon CH, Chung NS, Kwon HJ. Comparative Analysis of Three Imaging Modalities for Evaluation of Cervical Sagittal Alignment Parameters: A Validity and Reliability Study. Spine (Phila Pa 1976). 2017;42(24):1901–7. doi:10.1097/BRS.0000000000002256.

57. Li G, Passias P, Kozanek M, Shannon BD, Li G, Villamil F, et al. Interobserver reliability and intra-observer reproducibility of powers ratio for assessment of atlanto-occipital junction: comparison of plain radiography and computed tomography. Eur Spine J. 2009;18(4):577–82. doi:10.1007/s00586-008-0877-5.

58. Lopez WY, Goh BC, Upadhyaya S, Ziino C, Georgakas PJ, Gupta A, et al. Laminoplasty-an underutilized procedure for cervical spondylotic myelopathy. Spine J. 2021;21(4):571–7. doi:10.1016/j.spinee.2020.10.021.

59. Manabe A, Ishida T, Kanda E, Ono T. Evaluation of maxillary and mandibular growth patterns with cephalometric analysis based on cervical vertebral maturation: A Japanese cross-sectional study. PLoS One. 2022;17(4):e0265272. doi:10.1371/journal.pone.0265272.

60. Marshall TJ, Flower CD, Jackson JE. The role of radiology in the investigation and management of patients with haemoptysis. Clin Radiol. 1996;51(6):391–400. doi:10.1016/s0009-9260(96)80156-5.

61. Nakamura N, Inaba Y, Oba M, Aota Y, Morikawa Y, Ata Y, et al. Novel 2 radiographical measurements for atlantoaxial instability in children with Down syndrome. Spine (Phila Pa 1976). 2014 Dec 15;39(26):E1566–74. doi:10.1097/BRS.0000000000000625.

62. Paholpak P, Tamai K, Shoell K, Sessumpun K, Buser Z, Wang JC. Can multi-positional magnetic resonance imaging be used to evaluate angular parameters in cervical spine? A comparison of multi-positional MRI to dynamic plain radiograph. Eur Spine J. 2018;27(5):1021–7. doi:10.1007/s00586-017-5306-1.

63. Phillips FM, Phillips CS, Wetzel FT, Gelinas C. Occipitocervical neutral position. Possible surgical implications. Spine (Phila Pa 1976). 1999;24(8):775–8. doi:10.1097/00007632-199904150-00008.

64. Plaugher G, Cremata EE, Phillips RB. A retrospective consecutive case analysis of pretreatment and comparative static radiological parameters following chiropractic adjustments. J Manipulative Physiol Ther. 1990 Nov-Dec;13(9):498-506.

65. Rochester RP. Inter-and intra-examiner reliability of the upper cervical x-ray marking system: A third and expanded look. Chiropr Res J. 1994;3(1):23–31.

66. Salameh M, Bizdikian AJ, Saad E, Saliby RM, Nacouzi R, Khalil N, et al. Reliability assessment of cervical spine parameters measured on full-body radiographs in asymptomatic subjects and patients with spinal deformity. Orthop Traumatol Surg Res. 2021 Nov;107(7):103026. doi: 10.1016/j.otsr.2021.103026.

67. Shin JK, Lee JS, Kang SS, Lee JM, Youn BH. The reliabilities of radiographic measurements of cervical sagittal alignment in ankylosing spondylitis. Clin Spine Surg. 2016 Jul;29(6):E282–7. doi: 10.1097/BSD.0000000000000105.

68. Shoda N, Takeshita K, Seichi A, Akune T, Nakajima S, Anamizu Y, et al. Measurement of occipitocervical angle. Spine (Phila Pa 1976). 2004 May 15;29(10):E204-8. doi: 10.1097/00007632-200405150-00022.

69. Sigler DC, Howe JW. Inter- and intra-examiner reliability of the upper cervical X-ray marking system. J Manipulative Physiol Ther. 1985 Jun;8(2):75–80. PMID: 4009070.

70. Silber JS, Lipetz JS, Hayes VM, Lonner BS. Measurement variability in the assessment of sagittal alignment of the cervical spine: a comparison of the gore and cobb methods. J Spinal Disord Tech. 2004 Aug;17(4):301–5. doi: 10.1097/01.bsd.0000095824.98982.53.

71. Svedmark P, Lundh F, Németh G, Noz ME, Maguire GQ Jr, Zeleznik MP, et al. Motion analysis of total cervical disc replacements using computed tomography: preliminary experience with nine patients and a model. Acta Radiol. 2011 Dec;52(10):1128–37. doi: 10.1258/ar.2011.110230.

72. Tang C, Yang S, Liao YH, Tang Q, Ma F, Wang Q, et al. A novel method for measurement of the occipital-cervical distance via the occiput-C4 distance. BMC Musculoskelet Disord. 2020 Jun 15;21(1):385. doi: 10.1186/s12891-020-03398-9.

73. Vidal C, Ilharreborde B, Azoulay R, Sebag G, Mazda K. Reliability of cervical lordosis and global sagittal spinal balance measurements in adolescent idiopathic scoliosis. Eur Spine J. 2013 Jun;22(6):1362–7. doi: 10.1007/s00586-013-2752-2.

74. Wellborn CC, Sturm PF, Hatch RS, Bomze SR, Jablonski K. Intraobserver reproducibility and interobserver reliability of cervical spine measurements. J Pediatr Orthop. 2000 Jan-Feb;20(1):66-70.

75. Wu SK, Jou JY, Lee HM, Chen HY, Su FC, Kuo LC. The reproducibility comparison of two intervertebral translation measurements in cervical flexion-extension. Spine J. 2015 May;15(5):1083–91. doi: 10.1016/j.spinee.2013.06.097.

76. Zhang J, Zhang C, Zhong W, et al. Validity and reliability of a novel iPhone method to rapidly measure cervical sagittal parameters. Sci Rep. 2022;12:19579. doi: 10.1038/s41598-022-21660-z.

77. Ranganathan P, Pramesh CS, Aggarwal R. Common pitfalls in statistical analysis: Measures of agreement. Perspect Clin Res. 2017 Oct-Dec;8(4):187-91. doi: 10.4103/picr.PICR_123_17.

78. Alfuth M, Fichter P, Knicker A. Leg length discrepancy: A systematic review on the validity and reliability of clinical assessments and imaging diagnostics used in clinical practice. PLoS One. 2021;16:e0261457. doi: 10.1371/journal.pone.0261457.

79. Konieczka C, Gibson C, Russett L, Dlot L, MacDermid J, Watson L, et al. What is the reliability of clinical measurement tests for humeral head position? A systematic review. J Hand Ther. 2017;30(4):420–31. doi: 10.1016/j.jht.2017.06.010.

80. Dalton JE, Bolen SD, Mascha EJ. Publication bias: the elephant in the review. Anesth Analg. 2016 Oct;123(4):812–3. doi: 10.1213/ANE.0000000000001596.

81. Stoll CRT, Izadi S, Fowler S, Green P, Suls J, Colditz GA. The value of a second reviewer for study selection in systematic reviews. Res Synth Methods. 2019;10(4):539–45. doi: 10.1002/jrsm.1369.

82. Campbell M, McKenzie JE, Sowden A, Katikireddi SV, Brennan SE, Ellis S, et al. Synthesis without meta-analysis (SWiM) in systematic reviews: reporting guideline. BMJ. 2020;368:l6890. doi: 10.1136/bmj.l6890.

83. Barnett I, Malik N, Kuijjer ML, Mucha PJ, Onnela JP. Endnote: feature-based classification of networks. Netw Sci. 2019;7(4):438–44. doi: 10.1017/nws.2019.21.

84. Corso M, Cancelliere C, Mior S, Kumar V, Smith A, Côté P. The clinical utility of routine spinal radiographs by chiropractors: a rapid review of the literature. Chiropr Man Therap. 2020 Jul 9;28(1):33. doi: 10.1186/s12998-020-00323-8.

85. Williams B, Gichard L, Johnson D, Louis M. An investigation into the chiropractic practice and communication of routine, repetitive radiographic imaging for the location of postural misalignments. J Clin Imaging Sci. 2024;14:28. doi: 10.25259/JCIS_68_2024.

86. Bussieres AE, Ammendolia C, Peterson C, Taylor JA. Ionizing radiation exposure—more good than harm? The preponderance of evidence does not support abandoning current standards and regulations. J Can Chiropr Assoc. 2006;50(2):103–6.

87. Bussieres AE, Taylor JA, Peterson C. Diagnostic imaging practice guidelines for musculoskeletal complaints in adults—an evidence-based approach—part 3: spinal disorders. J Manipulative Physiol Ther. 2008;31(1):33–88. doi: 10.1016/j.jmpt.2007.11.003.

88. Jenkins HJ, Downie AS, Moore CS, French SD. Current evidence for spinal X-ray use in the chiropractic profession: a narrative review. Chiropr Man Therap. 2018;26:48. doi: 10.1186/s12998-018-0217-8.

89. Oakley PA, Moustafa IM, Haas JW, Betz JW, Harrison DE. Two methods of forward head posture assessment: radiography vs. posture and their clinical comparison. J Clin Med. 2024;13(7):2149. doi: 10.3390/jcm13072149.

90. Hosseini MM, Mahoor MH, Haas JW, Ferrantelli JR, Dupuis AL, Jaeger JO, et al. Intra-examiner reliability and validity of sagittal cervical spine mensuration methods using deep convolutional neural networks. J Clin Med. 2024;13(9):2573. doi: 10.3390/jcm13092573.

91. Haas JW, Oakley PA, Ferrantelli JR, Katz EA, Moustafa IM, Harrison DE. Abnormal static sagittal cervical curvatures following motor vehicle collisions: a retrospective case series of 41 patients before and after a crash exposure. Diagnostics (Basel). 2024;14(9):957. doi: 10.3390/diagnostics14090957.

92. Lippa L, Lippa L, Cacciola F. Loss of cervical lordosis: what is the prognosis? J Craniovertebr Junction Spine. 2017 Jan-Mar;8(1):9-14. doi: 10.4103/0974-8237.199877.

93. Suzuki H, Funaba M, Fujimoto K, Ichihara Y, Nishida N, Sakai T. Current concepts of cervical spine alignment, sagittal deformity, and cervical spine surgery. J Clin Med. 2024;13(5):1196. doi: 10.3390/jcm13051196.

94. Canseco JA, Karamian BA, Patel PD, Markowitz M, Lee JK, Kurd MF, et al. Perioperative change in cervical lordosis and health-related quality-of-life outcomes. Int J Spine Surg. 2022;16(6):960–8. doi: 10.14444/8325.

95. Romani MD, Zhang HQ, Gao QL, Liu SH, Deng A. Cervical sagittal alignment and related factor analysis and prediction model in patients undergoing revision surgery after anterior cervical fusion. J Am Acad Orthop Surg. 2024;32(12):e585–e595. doi: 10.5435/JAAOS-D-23-00565.

96. Figas G, Kostka J, Pikala M, Kujawa JE, Adamczewski T. Analysis of clinical pattern of musculoskeletal disorders in the cervical and cervico-thoracic regions of the spine. J Clin Med. 2024;13(3):840. doi: 10.3390/jcm13030840.

97. Moustafa IM, Ozsahin DU, Mustapha MT, Ahbouch A, Oakley PA, Harrison DE. Utilizing machine learning to predict post-treatment outcomes in chronic non-specific neck pain patients undergoing cervical extension traction. Sci Rep. 2024;14(1):11781. doi: 10.1038/s41598-024-62812-7.

98. Ma X, Yu Z, Wu D, Huang Y. Comparative analysis of postoperative sagittal balance in expansive open-door laminoplasty versus laminectomy with fusion for multilevel ossification of posterior longitudinal ligament: a retrospective study. Med Sci Monit. 2024;30:e943057. doi: 10.12659/MSM.943057.

99. Shi L. The impact of primary care: a focused review. Scientifica (Cairo). 2012;2012:432892. doi: 10.6064/2012/432892.

100. Oakley PA, Betz JW, Harrison DE, Siskin LA, Hirsh DW. Radiophobia overreaction: College of chiropractors of British Columbia revoke full x-ray rights based on flawed study and radiation fear-mongering. Dose-Response. 2021;19(3):15593258211033142. doi: 10.1177/15593258211033142.

101. Jung JY, Lee YB, Kang CK. Effect of forward head posture on resting state brain function. Healthcare (Basel). 2024;12(12):1162. doi: 10.3390/healthcare12121162.

102. Szczygieł E, Fudacz N, Golec J, Golec E. The impact of the position of the head on the functioning of the human body: a systematic review. Int J Occup Med Environ Health. 2020;33(5):559–68. doi: 10.13075/ijomeh.1896.01585.

103. Shahidi B, Haight A, Maluf K. Differential effects of mental concentration and acute psychosocial stress on cervical muscle activity and posture. J Electromyogr Kinesiol. 2013;23(4):1082–9. doi: 10.1016/j.jelekin.2013.05.009.

104. Mahmoud NF, Hassan KA, Abdelmajeed SF, Moustafa IM, Silva AG. The relationship between forward head posture and neck pain: A systematic review and meta-analysis. Curr Rev Musculoskelet Med. 2019;12:562–77. doi:10.1007/s12178-019-09594-y.

105. Katz EA, Katz SB, Fedorchuk CA, Lightstone DF, Banach CJ, Podoll JD. Increase in cerebral blood flow indicated by increased cerebral arterial area and pixel intensity on brain magnetic resonance angiogram following correction of cervical lordosis. Brain Circ. 2019;5(1):19–26. doi:10.4103/bc.bc_25_18.

106. Bulut MD, Alpayci M, Şenköy E, Bora A, Yazmalar L, Yavuz A, et al. Decreased vertebral artery hemodynamics in patients with loss of cervical lordosis. Med Sci Monit. 2016;22:495–500. doi:10.12659/MSM.897500.

107. Goo BW, Oh JH, Kim JS, Lee MY. Effects of cervical stabilization with visual feedback on craniovertebral angle and proprioception for the subjects with forward head posture. Medicine (Baltimore). 2024;103(2):e36845. doi:10.1097/MD.0000000000036845.

108. Titcomb DA, Melton BF, Bland HW, Miyashita T. Evaluation of the Craniovertebral Angle in Standing versus Sitting Positions in Young Adults with and without Severe Forward Head Posture. Int J Exerc Sci. 2024;17(1):73–85. doi:10.70252/GDNN4363.

109. Koseki T, Kakizaki F, Hayashi S, Nishida N, Itoh M. Effect of forward head posture on thoracic shape and respiratory function. J Phys Ther Sci. 2019;31(1):63–8. doi:10.1589/jpts.31.63.

110. Behzadi F, Luy DD, Zsigray B, Uram Z, Iordanou J, Ng IB, et al. Posterior Circulation Ischemic Stroke Is Associated with More Severe Forward Head Posture in Patients with Cervicalgia. World Neurosurg. 2024;190:e570–8. doi:10.1016/j.wneu.2024.07.178.

111. Oakley PA, Ehsani NN, Moustafa IM, Harrison DE. Restoring cervical lordosis by cervical extension traction methods in the treatment of cervical spine disorders: a systematic review of controlled trials. J Phys Ther Sci. 2021;33(10):784–94. doi:10.1589/jpts.33.784.

112. Patwardhan AG, Khayatzadeh S, Havey RM, Voronov LI, Smith ZA, Kalmanson O, et al. Cervical sagittal balance: a biomechanical perspective can help clinical practice. Eur Spine J. 2018;27(Suppl 1):25–38. doi:10.1007/s00586-017-5367-1.

113. Moustafa IM, Diab AA, Hegazy F, Harrison DE. Demonstration of central conduction time and neuroplastic changes after cervical lordosis rehabilitation in asymptomatic subjects: a randomized, placebo-controlled trial. Sci Rep. 2021;11(1):15379. doi:10.1038/s41598-021-94548-z.

114. Norton TC, Oakley PA, Haas JW, Harrison DE. Positive Outcomes Following Cervical Acceleration-Deceleration (CAD) Injury Using Chiropractic BioPhysics® Methods: A Pre-Auto Injury and Post-Auto Injury Case Series. J Clin Med. 2023;12(19):6414. doi:10.3390/jcm12196414.

115. Cheng J, Liu P, Sun D, Ma Z, Liu J, Wang Z, et al. Correlation of cervical and thoracic inlet sagittal parameters by MRI and radiography in patients with cervical spondylosis. Medicine (Baltimore). 2019;98(7):e14393. doi:10.1097/MD.0000000000014393.

116. Zhang Z, Wang J, Ge R, Guo C, Liang Y, Liu H, et al. A novel classification that defines the normal cervical spine: an analysis based on 632 asymptomatic Chinese volunteers. Eur Spine J. 2024;33(1):155–65. doi:10.1007/s00586-023-07997-7.

117. Mekhael E, El Rachkidi R, Saliby RM, Nassim N, Semaan K, Massaad A, et al. Functional assessment using 3D movement analysis can better predict health-related quality of life outcomes in patients with adult spinal deformity: a machine learning approach. Front Surg. 2023;10:1166734. doi:10.3389/fsurg.2023.1166734.

118. Javaid M, Haleem A, Pratap Singh R, Suman R, Rab S. Significance of Machine Learning in Healthcare: Features, Pillars and Applications. Int J Intell Netw. 2022;3:58–73. doi:10.1016/J.IJIN.2022.05.002.

119. Zorina-Lichtenwalter K, Bango CI, Van Oudenhove L, Čeko M, Lindquist MA, Grotzinger AD, et al. Genetic risk shared across 24 chronic pain conditions: identification and characterization with genomic structural equation modeling. Pain. 2023;164(10):2239–52. doi:10.1097/j.pain.0000000000002922.

120. Kang KC, Lee HS, Lee JH. Cervical radiculopathy focus on characteristics and differential diagnosis. Asian Spine J. 2020;14(6):921–30. doi:10.31616/asj.2020.0647.

121. Kotecki M, Gasik R, Głuszko P, Sudoł-Szopińska I. Radiological evaluation of cervical spine involvement in rheumatoid arthritis: A cross-sectional retrospective study. J Clin Med. 2021;10(19):4587. doi:10.3390/jcm10194587.

122. Guo GM, Li J, Diao QX, Zhu TH, Song ZX, Guo YY, et al. Cervical lordosis in asymptomatic individuals: a meta-analysis. J Orthop Surg Res. 2018;13(1):147. doi:10.1186/s13018-018-0854-6.

123. Kim KH, Park SA, An CS, Kim JL. The relationship between cervical lordosis and neck pain: A systematic review and meta-analysis. Appl Sci Tech. 2024;8(5):1666–75. doi:10.55214/25768484.v8i5.1885.

124. Harrison DD, Janik TJ, Troyanovich SJ, Holland B. Comparisons of lordotic cervical spine curvatures to a theoretical ideal model of the static sagittal cervical spine. Spine (Phila Pa 1976). 1996;21(6):667–75. doi:10.1097/00007632-199603150-00002.

125. Harrison DD, Harrison DE, Janik TJ, Cailliet R, Ferrantelli JR, Haas JW, et al. Modeling of the sagittal cervical spine as a method to discriminate hypolordosis: results of elliptical and circular modeling in 72 asymptomatic subjects, 52 acute neck pain subjects, and 70 chronic neck pain subjects. Spine (Phila Pa 1976). 2004;29(22):2485–92. doi:10.1097/01.brs.0000144449.90741.7c.

126. Yen CY, Lin SM, Chen HY, Wang SW, Tsai YD, Chye CL, et al. Sagittal alignment to predict efficiency in pulsed radiofrequency for cervical facet joint pain. Sci Rep. 2024;14(1):28563. doi:10.1038/s41598-024-79181-w.

127. Cao Y, Xu C, Sun B, Cui C, Zhang K, Wu H, et al. Preoperative Cervical Cobb Angle Is a Risk Factor for Postoperative Axial Neck Pain after Anterior Cervical Discectomy and Fusion with Zero-Profile Interbody. Orthop Surg. 2022;14(12):3225–32. doi:10.1111/os.13552.

128. Liang CL, Wang SW, Chen HJ, Tsai YD, Chen JS, Wang HK, et al. Optimal Cut-Off Points of Sagittal Spinopelvic Parameters as a Morphological Parameter to Predict Efficiency in Nerve Block and Pulsed Radiofrequency for Lumbar Facet Joint Pain: A Retrospective Study. J Pain Res. 2021;14:1949–57. doi:10.2147/JPR.S303979.

129. Viswanathan M, Patnode CD, Berkman ND, et al. Assessing the Risk of Bias in Systematic Reviews of Health Care Interventions. In: Methods Guide for Effectiveness and Comparative Effectiveness Reviews [Internet]. Rockville (MD): Agency for Healthcare Research and Quality (US); 2008–. Available from: https://www.ncbi.nlm.nih.gov/books/NBK519366/

